# Aberrant transcript usage induces homologous recombination deficiency and predicts therapeutic responses

**DOI:** 10.1101/2021.08.31.21262939

**Authors:** Hyeon Gu Kang, Haeun Hwangbo, Myung Ji Kim, Sinae Kim, Eun Ji Lee, Min Ji Park, Jae-Weon Kim, Byoung-Gie Kim, Eun-Hae Cho, Suhwan Chang, Jung-Yun Lee, Jung Kyoon Choi

**Author notes:** Correspondence: SC, EHC, JYL, and JKC.

## Abstract

BRCA1/2 mutations account for only a small fraction of homologous recombination (HR) deficiency (HRD) cases. Recently developed genomic HRD (gHRD) tests suffer confounding factors causing low precision in predicting samples that will respond to poly (ADP-ribose) polymerase (PARP) inhibitors and DNA damaging agents. Here, we present molecular evidence and clinical utility of transcriptional HRD (tHRD) that is based on aberrant transcript usage (TU) of minor isoforms. Specifically, increased TU of non-functional isoforms of DNA repair genes was prevalent in breast and ovarian cancer with gHRD. Our functional assays validated its association with impaired HR activity. Remarkably, tHRD detection based on the TU pattern of key genes was superior to gHRD or BRCA1/2 screening in accuracy for predicting the responses of cell lines and cancer patients to PARP inhibitors and genotoxic drugs. In particular, this approach demonstrated the capability to reflect functional HR status, particularly when applied to our cohort of olaparib users with acquired platinum resistance in ovarian cancer. Hence, the tHRD-based diagnostic tests are expected to broaden the clinical utility of PARP inhibitors.

## INTRODUCTION

The two key genes of the homologous recombination (HR) pathway for double-strand break (DSB) repair, BRCA1 and BRCA2, are intimately associated with the heritability of breast and ovarian cancer. Recent studies have shown that DNA damage response pathways frequently are mutated in these and other cancers^1–3^. Therefore, targeting vulnerabilities in DNA repair activity, in particular HR deficiency (HRD), has been a primary treatment option. In the representative example of synthetic lethality, inhibiting poly (ADP-ribose) polymerase (PARP) enzymes can selectively suppress cells that are defective in DNA damage responses, for example, due to BRAC1/2 mutations^4–9^.

However, BRCA1/2 mutations account for a small fraction of HRD cases, thereby limiting the development of more comprehensive HRD diagnostic markers. A promising approach has been to detect the genomic consequences, instead of the causal factors, of HRD. Defective HR machinery leaves genomic footprints and lesions as a consequence of failure in damage maintenance. Specifically, signature 3 and genomic scar are well known indicators of HRD^10–15^. Signature 3 has been linked to HRD by associating the genome-wide patterns of single nucleotide variants with specific background factors^16^. Genomic scar is determined by a combination of three chromosomal aberrant events, namely, telomeric allelic imbalances, large-scale state transition, and loss of heterozygosity^17–19^.

These genomic assays, however, cannot reflect the functional restoration of HR that gives rise to therapeutic resistance^20^. BRCA1/2 reversion and various other mechanisms can account for acquired resistance to platinum regimens and PARP inhibitors in ovarian cancer^21–27^. In these cases, initially imprinted genomic scars remain detectable in tumors with recovered HR activity. Furthermore, several technical artifacts can arise when calculating mutational signatures^28^. For example, the bleeding of signatures refers to the phenomenon in which signatures present in only some samples of a cohort affect the signature assignment of the entire cohort. Directly related to detection of HRD is the tendency of the fitting processes to force the assignment of flat signature 3 to irrelevant samples^28^.

These biological or technical confounding factors together make detection of genomic HRD (gHRD) prone to false positives. This results in low levels of precision because a considerable number of tumors predicted to respond to DNA damaging agents or PARP inhibitors would not actually respond^29^. Because of the coupling of precision and recall, attempts to improve precision by applying more stringent cutoffs will undermine sensitivity. This calls for a different biomarker that can complement this limitation and improve precision. For example, biomarkers that can detect functional HRD status may be used to refine gHRD predictions by filtering out tumors that regained HR functionality^29^.

In this work, we pay attention to transcriptional processes, which change dynamically in response to functional fluctuation during cancer evolution but are not affected significantly by the genomic consequences of HRD. We employ machine learning to capture patterns of transcriptional aberration that may be responsible for true gHRD readouts by contrasting with the profiles of negative gHRD samples. Ultimately, by leveraging data from patients with acquired resistance to initial treatments, we attempt to test whether our method can determine real-time, functional HR status and overcome the limitations imposed by the artefacts of the gHRD tests.

## RESULTS AND DISCUSSION

### Minor isoform overexpression is associated with gHRD

We collected breast cancer samples from The Cancer Genome Atlas (TCGA), examined germline and somatic BRCA1/2 mutations, and defined HR status by using both genomic scar and signature 3 readouts (Supplementary Fig. 1A). As expected, a majority (> 87%) of BRCA1/2-mutant samples were gHRD+ (Supplementary Fig. 1B, left). Importantly, a sizeable fraction (∼ 44%) of BRCA1/2-wildtype tumors also showed genomic instability at a stringent threshold, satisfying both metrics (Supplementary Fig. 1B, right), thereby implying HRD mechanisms that do not engage BRCA1/2 mutations. By using the transcriptomes of TCGA samples, we tested whether DNA repair genes were transcriptionally inactive in the gHRD+ samples. However, the genes repressed in association with gHRD+ were not enriched for molecular functions associated with DNA repair (Supplementary Fig. 1C).

This prompted us to investigate differential RNA splicing between the gHRD+ and gHRD− samples. Remarkably, our enrichment analysis indicated that genes involved in DNA repair tend to be spliced differentially between the two groups (Supplementary Fig. 2A). However, splicing patterns themselves are not indicative of functional consequences. To further test whether splicing regulation is responsible for HRD, we examined the patterns of transcript usage (TU) because minor or alternative isoforms can directly represent functional loss^30, 31^. Specifically, we identified aberrant transcript usage (aTU) as the relative overexpression of a minor transcript in the gHRD+ samples (Fig. 1A, B). In total, we identified 2,521 such aTU genes (Fig. 1B) with a significant overrepresentation of DNA repair-related terms (Supplementary Fig. 2B), particularly DSB repair and DNA replication (red dots in Fig. 1C). The aTU patterns of representative DSB repair genes, including RAD51, are shown in Fig. 1D. Notably, this enrichment was not found for the opposite definition of aTU with minor isoform overexpression in gHRD− tumors (blue dots in Fig. 1C).

**Figure 1.**
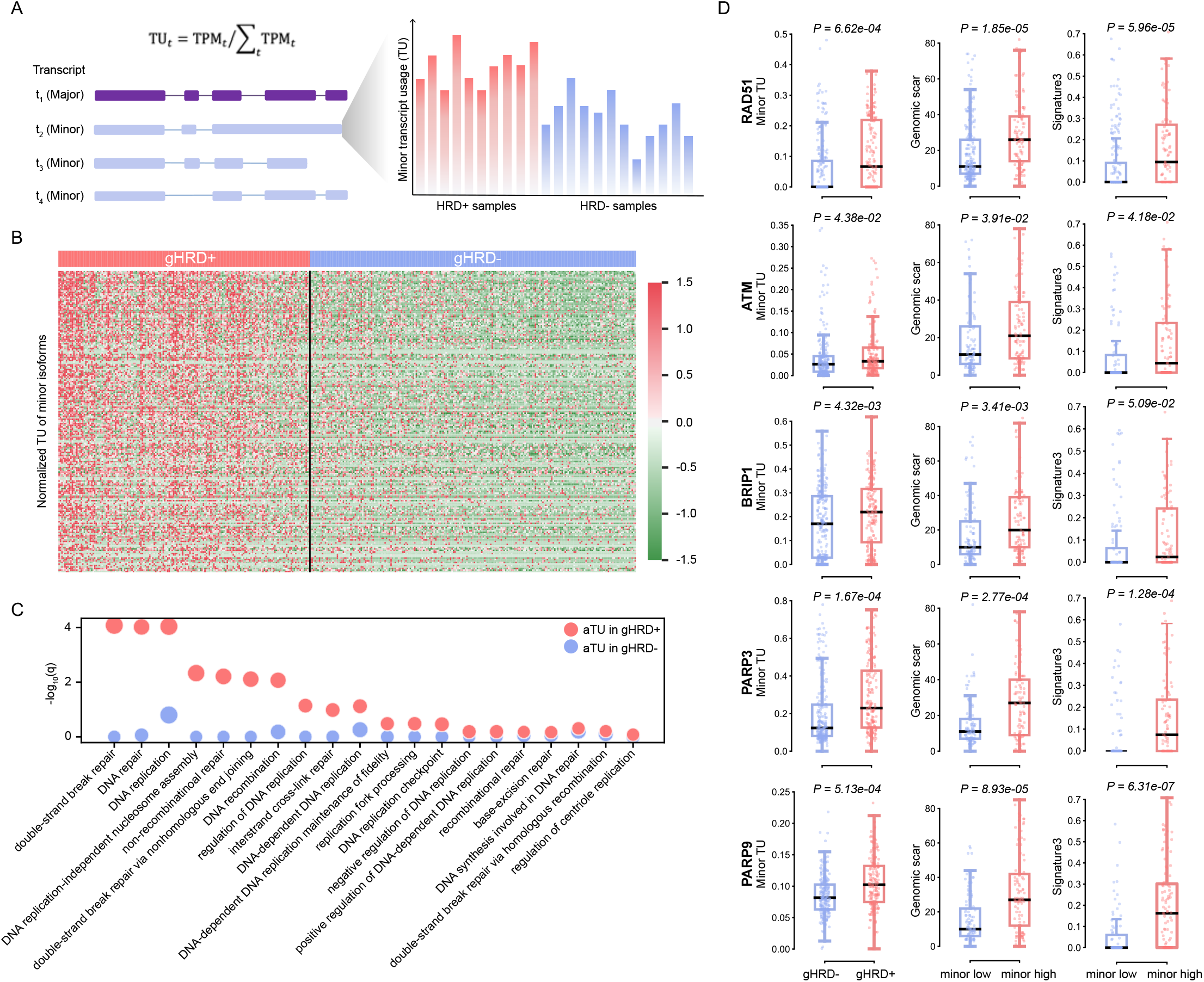
HRD-associated overexpression of minor transcripts of DNA repair genes. **(A)** Concept of aTU as the relative overexpression of a minor isoform of a gene in HRD+ tumors. TU is calculated as the transcripts per million (TPM) of an isoform divided by the sum of all TPMs. From each gene, the minor isoform with the most significant differential TU was selected and used to calculate the representative minor TU of the given gene. **(B)** Normalized minor TU levels of 2,521 aTU genes in gHRD+ versus gHRD− TCGA breast tumors. Significantly differential TU was identified at a false discovery rate < 10%. **(C)** Functional enrichment analysis of the 2,521 aTU genes (red dots) in comparison to a negative control set for which aTU was defined as the relative overexpression of a minor isoform in gHRD− (blue dots). **(D)** Representative aTU genes involved in DSB repair. Shown are the levels of the minor TU of the gene in gHRD+ versus gHRD− samples (left), genomic scar in samples with high minor TU versus low minor TU (middle), and signature 3 in samples with high minor TU versus low minor TU (right). High and low minor TU are defined at the 75^th^ and 25^th^ percentile of all samples, respectively.

We examined whether the gHRD-associated aTU events give rise to functional loss. In fact, the frequency of retrained introns and processed transcripts (i.e., noncoding transcripts that do not contain an open reading frame) by aTU was >3 times higher than expected from the genome-wide distribution (Fig. 2A). Non-functional minor forms overexpressed in gHRD+ from ATM, BRIP1, RAD51, and MRE11 are shown in Fig. 2B. For example, intron retention in BRIP1 results in the disruption of the BRCA1-interacting domain by a frameshift (Fig. 2B). We checked the RNA read profiles of aTU involving intron retention in BRIP1 and other genes in selected samples (red for gHRD+ samples and blue for gHRD− samples in Fig. 2C). The aTU patterns were observed specifically in gHRD+ tumors but not in their matched normal samples (Supplementary Fig. 3).

**Figure 2.**
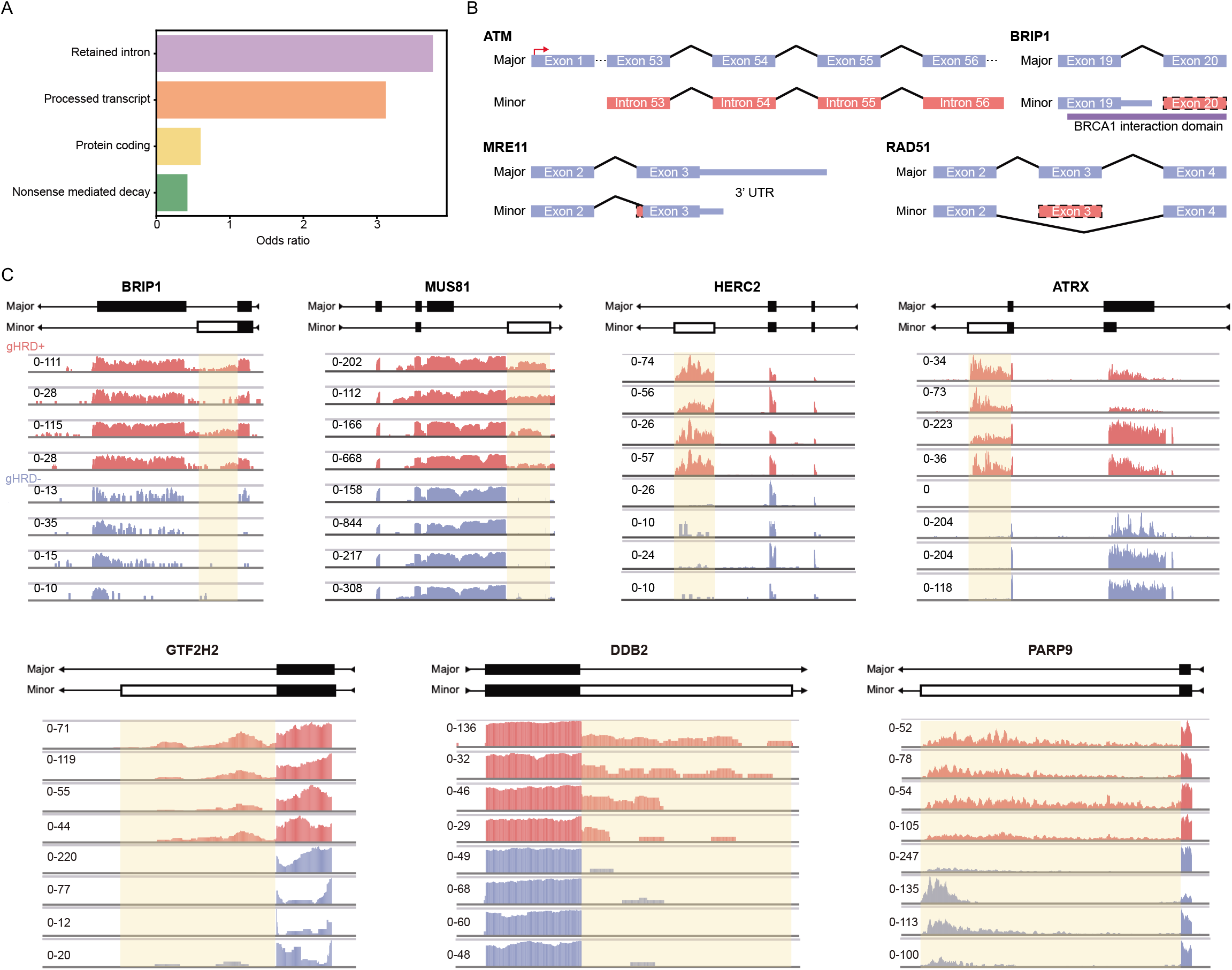
Functional characterization of aTU events. **(A)** Frequencies of annotation categories that represent the functional consequences or alternative events by minor isoform transcription. The odds of each class among the identified aTU genes was divided by the odds of that class among all genes. For example, a >3-fold overrepresentation of intron retention and noncoding transcription was found among minor isoforms overexpressed in gHRD+ relative to all such events in the genome. **(B)** Non-functional minor transcripts overexpressed in gHRD+ from ATM, BRIP1, MRE11, RAD51, and MRE11. **(C)** RNA read profiles of aTU involving intron retention in representative genes in selected samples (red for gHRD+ samples and blue for gHRD− samples.

### Development and functional validation of the tHRD detection model

We selected 35 DNA repair genes that showed aTU in TCGA breast cancer and developed a tHRD prediction model based on the TU level of their transcripts (Supplementary Fig. 4A and Supplementary Table 1). The performance of the aTU-based classifier in predicting gHRD indicates only the degree of agreement between the tHRD and gHRD methods. Some gHRD+ samples may be predicted as tHRD− when they present artefactual genomic signatures as described in the Introduction. Also, some gHRD− samples may be predicted to be tHRD+ when they share similar TU patterns with genuine gHRD+ samples; these should not be considered as false positives. Hence, the tHRD results should be validated by functional or clinical measures independently of gHRD status. For example, HRD+ tumors are expected to show poor prognosis and high mutation burden. In this regard, the tHRD approach was as successful as the gHRD method (Supplementary Fig. 4B).

To perform more detailed functional tests of the tHRD results in a gHRD-independent manner, we first employed the concept of genetic dependencies. Substantial effort has been put into identifying genes essential for cancer cell proliferation and survival based on systematic loss-of-function screens^32–41^. Synthetic lethality arises when inactivation of one gene (e.g., by BRAC1/2 mutations) increases dependencies on a related gene (e.g., PARP1) so that the simultaneous perturbation of the two genes results in cell death^7^. In this respect, the samples predicted to be HRD+ should be dependent on genes with DNA repair activity.

We first tested this concept by using cell lines for which both transcriptome and dependency data were available. Cell lines with higher tHRD scores represented higher average dependencies on DNA repair genes (Supplementary Fig. 5). For validation in clinical samples, we performed dependency prediction^42^ for TCGA tHRD+ and tHRD− samples, and then examined enrichment of the predicted dependencies for DNA repair genes. The dependencies of the tHRD+ tumors, but not the tHRD− tumors, showed a significant overrepresentation of genes involved in DNA repair pathways (red versus blue bars in Fig. 3A). Indeed, higher dependency scores were observed in tHRD+ than in tHRD− for many DNA repair genes, with the largest difference for PRKDC (Fig. 3B-C). As the main component of the nonhomologous end joining pathway, DNA-PKcs (encoded by PRKDC) is recruited to DSBs for DNA repair when HR is unavailable or defective^43^. Further studies established synthetic lethality between the HR pathway and DNA-PKcs^44–47^. We discovered many genes involved in DNA repair and related pathways showing positive correlations between minor TU and PRKDC dependency (Fig. 3D).

**Figure 3.**
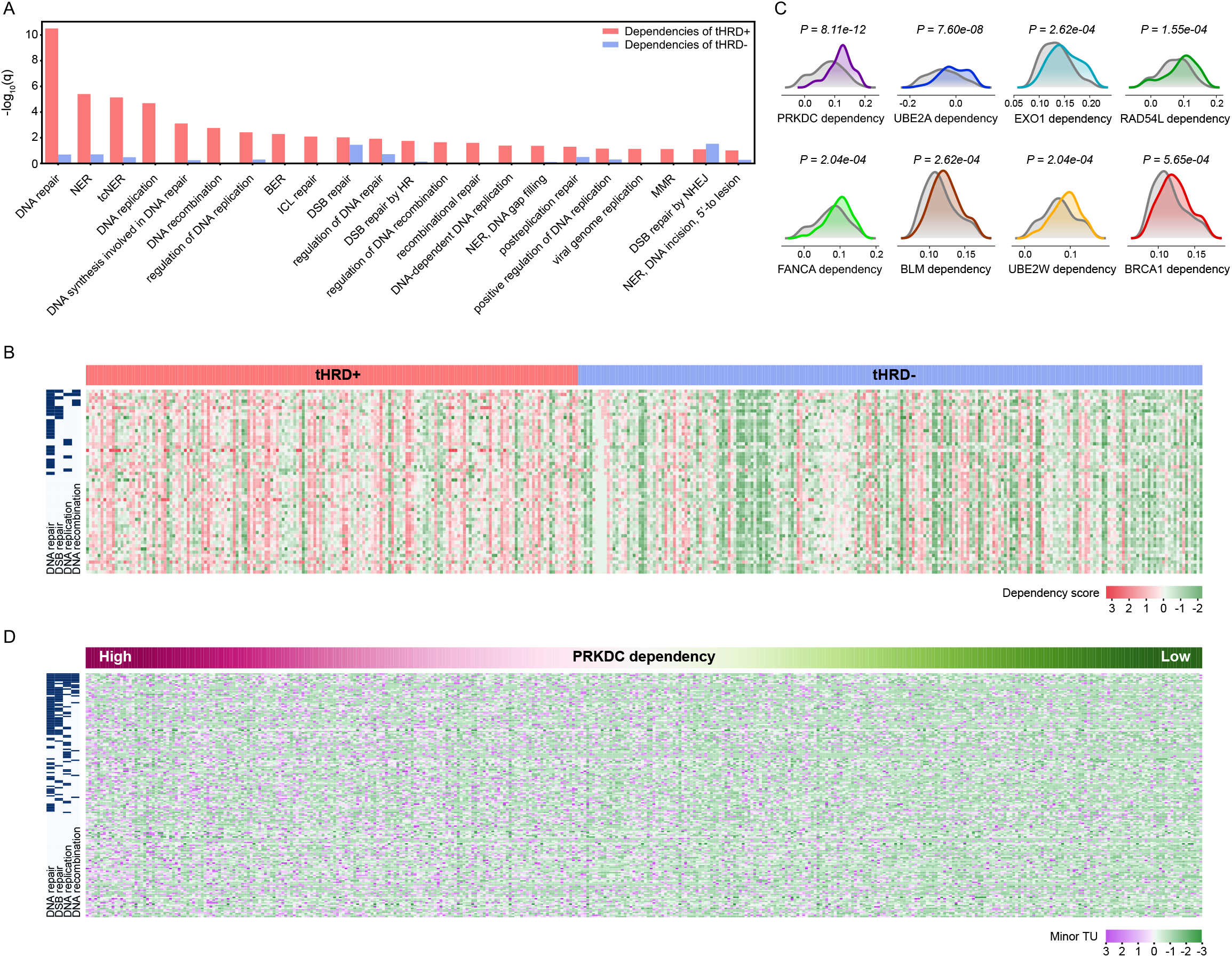
Functional validation of tHRD model based on genetic dependencies of clinical samples. **(A)** Functional enrichment analysis of the genes predicted to be dependencies in TCGA tHRD+ and tHRD− samples. Statistical significance of overrepresentation of the genes involved in various DNA repair pathways was plotted. **(B)** Higher dependency scores in TCGA tHRD+ and tHRD− samples (columns) were found for genes (rows) pertaining to DNA repair, recombination, and replication. Relevant functional categories are marked on the left side of the heatmap. Dependency scores for TCGA tumors were derived from our previous work^42^. **(C)** Examples of genes with higher dependency scores in the tHRD+ and tHRD− tumors. Colored and grey curves represent the distribution of the tHRD+ and tHRD− samples, respectively. **(D)** Genes (rows) whose minor isoform expression correlated with genetic dependencies on the PRKDC gene. Across TCGA samples (columns), we calculated the correlations between the PRKDC dependency score and the TU of minor isoforms of individual genes. Marked on the left are genes involved in the pathways of DNA repair, replication, and recombination.

We next sought to validate our tHRD prediction by functional HR assays in BRCA1/2-wildtype patient-derived (PD) cells. One tHRD+ sample (PD:tHRD+) and one tHRD− sample (PD:tHRD−) were selected by applying the tHRD model to the transcriptomes of our PD xenografts of breast cancer^48^. PD:tHRD+ overexpressed the minor isoforms of key repair genes, including RAD50, ATM, H2AX, and BRIP1 (Fig. 4A). In our first assay, DSB repair processes were visualized by γ-H2AX and RAD51 staining before and after olaparib treatment. Whereas olaparib treatment induced DSBs overall, a significantly larger number of γ-H2AX and RAD51 foci were observed in PD:tHRD+ than in PD:tHRD− (Fig. 4B and Supplementary Fig. 6A). For a more direct assessment of HR-mediated DSB repair activity, we performed the DR-GFP/I-*Sce*I HR assay^49^ in which GFP signals represent the full repair of introduced DSBs. As a result, PD:tHRD+ showed a 2∼3-fold lower fraction of GFP-positive cells than did PD:tHRD− (Fig. 4C and Supplementary Fig. 6B). These results collectively suggest that the accumulation of γ-H2AX/RAD51 foci in the PD:tHRD+ cells results from delayed DSB resolution due to impaired HR functionality, as demonstrated previously^50–53^.

**Figure 4.**
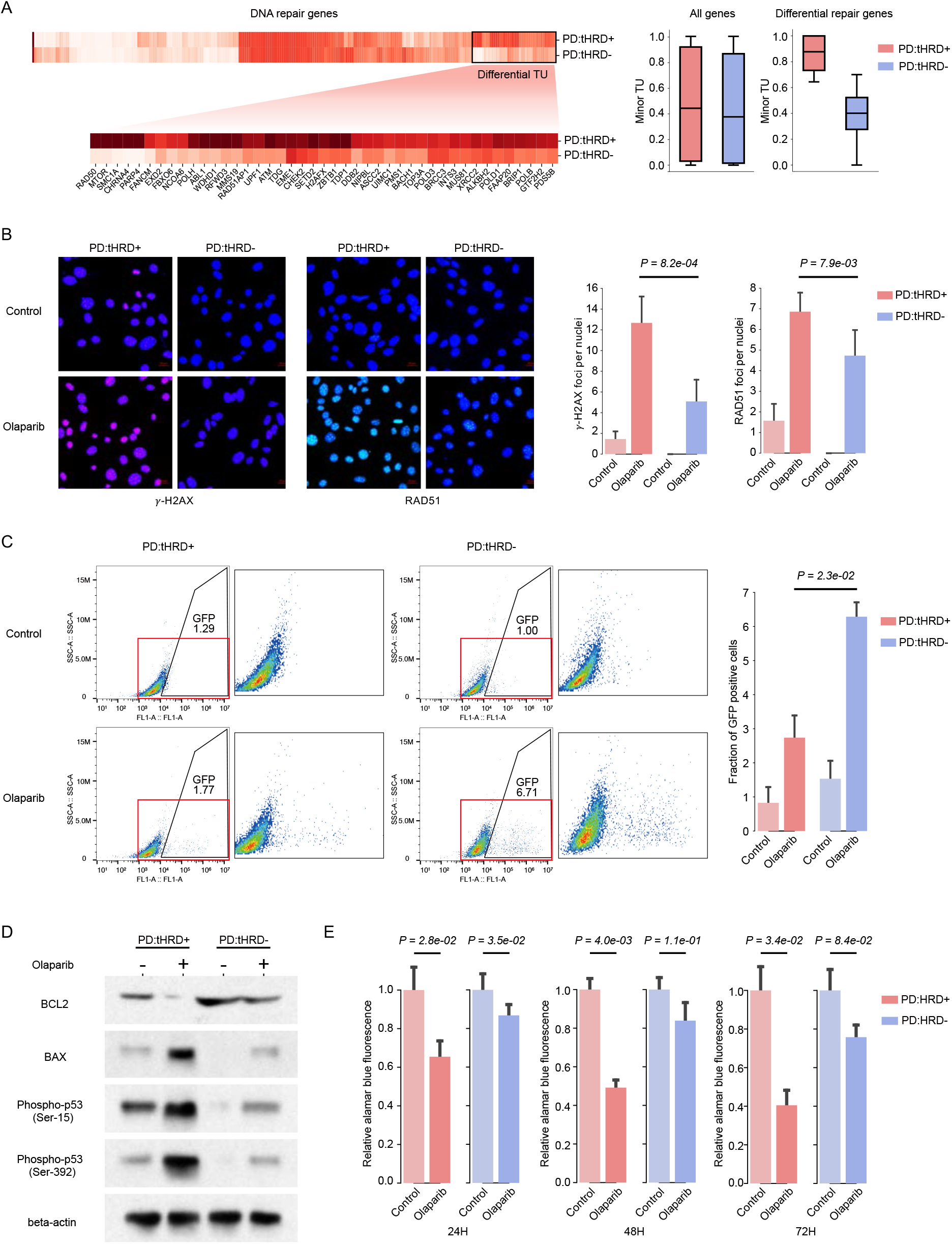
Functional validation of tHRD model by HR assays in olaparib-treated PD cells. **(A)** Comparison of the minor TU of DNA repair genes between PD tHRD+ cells (PD:tHRD+) and tHRD− cells (PD:tHRD−). A cluster of genes showing minor form overexpression in PD:tHRD+ was identified. Names of the genes are provided below the zoomed heatmap of differential TU. **(B)** Immunofluorescence staining images for γ-H2AX (red) and RAD51 (green) in PD:tHRD+ and PD:tHRD− before and after olaparib treatment. Blue background indicates the fluorescent stain DAPI. Additional images are provided in Supplementary Fig. 6A. The graphs show the number of γ-H2AX and Rad51 foci per cells with the mean and standard error obtained from four images. Comparisons between two groups were performed using the Student’s *t*-test. **(C)** Representative scatter plots showing the rate of HR repair based on the DR-GFP/I-*Sce*I assay in PD:tHRD+ and PD:tHRD− before and after olaparib treatment. GFP-positive cells in the marked and zoomed zones indicate a population of cells that underwent HR-mediated DSB repair of GFP reporter plasmids. The graph shows the fraction of the GFP-positive cells with the mean and standard error obtained from three replicate experiments after excluding the maximum and minimum. Comparisons between two groups were performed using the Student’s *t*-test. Replicate plots are provided in Supplementary Fig. 6B. **(D)** Western blot analysis of PD:tHRD+ and PD:tHRD− before and after olaparib treatment for BCL2 (antiapoptotic protein), BAX (proapoptotic protein), P53 phosphorylation at Ser-15 (senescence marker), and P53 phosphorylation at Ser-392 (apoptosis marker). **(E)** Cell viability measured by the alamarBlue® assay in PD:tHRD+ and PD:tHRD− before and after olaparib treatment. Fluorescence signals were obtained at 24, 48, and 72 hours after the treatment and normalized by the pre-treatment (control) measurements. Mean and standard error were obtained from three experiments. Comparisons between two groups were performed using the Student’s *t*-test.

To compare the cellular consequences of the differential rates of HR repair, we examined the phosphorylation of P53, the key player of DNA damage response. The phosphorylation of P53 at Ser-392 and at Ser-15 gives rise to apoptosis and cellular senescence, respectively^54, 55^. According to the Western blots, olaparib treatment strongly induced P53 phosphorylation, particularly at Ser-392 in PD:tHRD+ but not in PD:tHRD− (Fig. 4D). Accordingly, PD:tHRD+ presented an increase of the proapoptotic protein, BAX, and a decrease of the antiapoptotic protein, BCL2, in response to olaparib (Fig. 4D). These data suggest that the unresolved DSB foci of PD:tHRD+ (Fig. 4B-C) eventually lead to apoptosis of the cells. Indeed, our cell viability assays showed a significant reduction in cell proliferation when the PD:tHRD+ cells were challenged by olaparib (Fig. 4E).

### tHRD predicts drug responses with high accuracy in cell lines

To evaluate our tHRD model in predicting susceptibility to PARP inhibition or genotoxic stress, we examined the response of breast cancer cell lines to olaparib, rucaparib, talazoparib, veliparib, cisplatin, doxorubicin, bleomycin, etoposide, and SN-38 using the Genomics of Drug Sensitivity in Cancer (GDSC) database^56^. We applied our tHRD detection model to the RNA sequencing data of the cell lines to classify them as tHRD+ and tHRD−. (Supplementary Table 2). In terms of the half-maximal inhibitory concentration (IC_50_), significant differences were found in the sensitivity of the tHRD+ and tHRD− cell lines for all the PARP inhibitors and DNA damaging agents (Fig. 5A and Supplementary Fig. 7).

**Figure 5.**
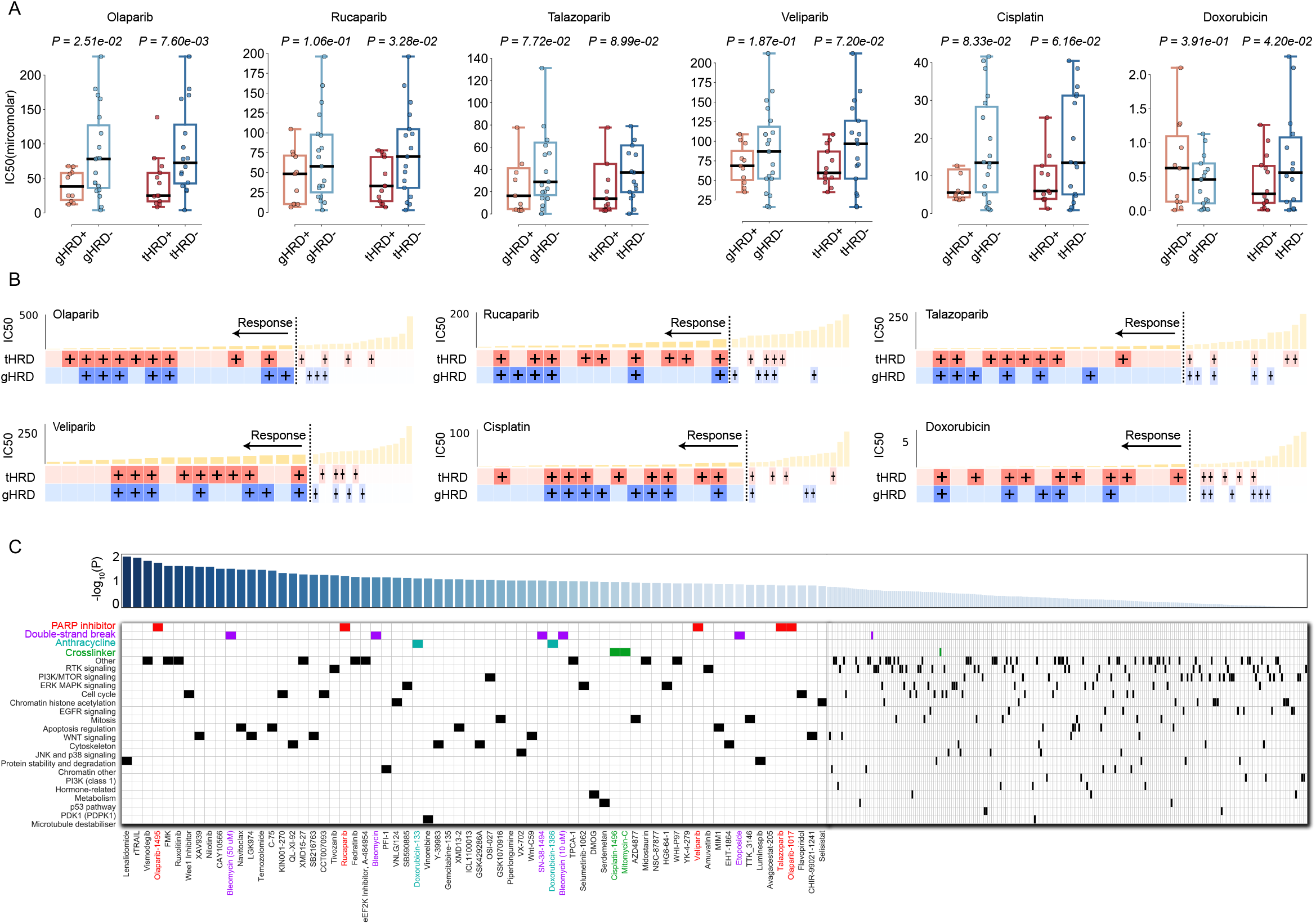
Responses to PARP inhibitors and genotoxic drugs explained by tHRD in cell lines. **(A)** Comparison of IC_50_ between tHRD+/tHRD− versus gHRD+/gHRD− in breast cancer cell lines. The same graphs for the responses to bleomycin, etoposide, and SN-38 are provided in Supplementary Fig. 7. Response data were obtained from the GDSC database. We applied our tHRD detection model to the RNA sequencing data of the cell lines to classify them as tHRD+ and tHRD−. **(B)** Mapping of drug sensitivity and HRD prediction results in respective cell lines. Below the plot of IC_50_ values are the results of our tHRD model and signature 3-based gHRD assay for the corresponding cell lines. The plus signs denote HRD+ predictions. gHRD was considered positive above the 75^th^ percentile of signature 3. IC_50_ values lower than the median were regarded as positive responses to the given drug. **(C)** Differences in drug sensitivity between the tHRD+ and tHRD− breast cancer cell lines across drug classes. Differences are denoted as −log_10_ P values at the top for drugs sorted in the order of the statistical significance. Below is the grouping of the drugs named at the bottom according to their mechanism of action shown on the left. PARP inhibitors, DSB-inducing agents, anthracyclines, and DNA cross-linkers are highlighted in colors.

Remarkably, the IC_50_ differences were more significant between the tHRD+ and tHRD− groups than between the gHRD+ and gHRD− groups (Fig. 5A) for almost all drugs, indicating that tHRD better predicts drug sensitivity than gHRD. For instance, the olaparib and rucaparib responses scored P = 0.008 (tHRD) vs P = 0.025 (gHRD) and P = 0.033 (tHRD) vs P = 0.106 (gHRD), respectively. We then examined the tHRD and gHRD status of individual samples. Notably, there were multiple cases in which the response was predicted only by tHRD but not by gHRD (i.e., tHRD+/gHRD− samples) in the responsive group (IC_50_ values lower than the median) (Fig. 5B). Lowering the threshold of gHRD (from the 75^th^ to 50^th^ percentile) led to many cases in which only gHRD made a positive prediction (i.e., gHRD+/tHRD− samples) in the no response group (IC_50_ values greater than the median) (Supplementary Fig. 8A). At this threshold, however, gHRD did not make a meaningful distinction between the positive and negative samples (see the P values above the boxplots of Supplementary Fig. 8B).

These data suggest that gHRD is prone to false positive signals attributed to biological or technical artefacts^29^. This feature of gHRD explains low levels of precision at a common cutoff of the median with a considerable number of the gHRD+ samples that do not actually respond to the drugs (Supplementary Fig. 8A). A more stringent threshold improves precision but sacrifices recall (also known as sensitivity), as demonstrated in Fig. 5B. In this case, the tHRD approach is more sensitive than the gHRD method and can identify tumors that do not harbor detectable genomic signatures but that respond to these drugs.

Additionally, we tested the drug specificity of the responses as reported previously^14^ by comparing IC_50_ among different classes of molecules grouped according to their mechanism of action. Specific responses were observed for PARP inhibitors (olaparib, rucaparib, veliparib, and talazoparib; red in Fig. 5C), DSB-inducing agents (bleomycin, SN-38, and etoposide; violet in Fig. 5C), and anthracyclines or DNA cross-linkers (doxorubicin, cisplatin, and mitomycin C; green in Fig. 5C).

### tHRD predicts drug responses with high accuracy in clinical samples

We next sought to assess the utility of tHRD in clinical settings. To first develop a tHRD model for ovarian cancer, the procedures we applied to TCGA breast tumors were repeated for TCGA ovarian samples; briefly, we partitioned the patient samples into gHRD+ and gHRD− groups, identified aTU cases with minor isoform overexpression in gHRD+, and developed an aTU-based prediction model with 22 DNA repair genes.

By using the reannotation of clinical outcomes of TCGA cases (n = 103)^57^, we evaluated the performance of our tHRD model in predicting platinum sensitivity (Supplementary Table 3). In comparison to gHRD, the tHRD prediction exhibited an improvement in terms of the precision and recall metrics as well as the accuracy and F1 score (Fig. 6A). Also, progression-free survival was substantially better explained by tHRD (P = 2.6×10^-6^) than gHRD (P = 4.7×10^-3^ or P = 1.1×10^-2^) (Fig. 6B). We further tested the model using samples from our neoadjuvant chemotherapy (NAC) cohort (n = 27) and performed validation by adding independent samples with platinum resistance (PR) (n = 36) (Supplementary Table 4). The tHRD model outperformed gHRD in the NAC cohort (Fig. 6C) and made no false positives in the PR cohort (Fig. 6D and Supplementary Fig. 9A). In contrast, gHRD made a substantial number of false positive predictions for the PR samples (Fig. 6D), failing to achieve a clinically applicable level of accuracy for the combined dataset (Supplementary Fig. 9A). BRCA1/2 testing was not successful at all in predicting platinum responses (Fig. 6D).

**Figure 6.**
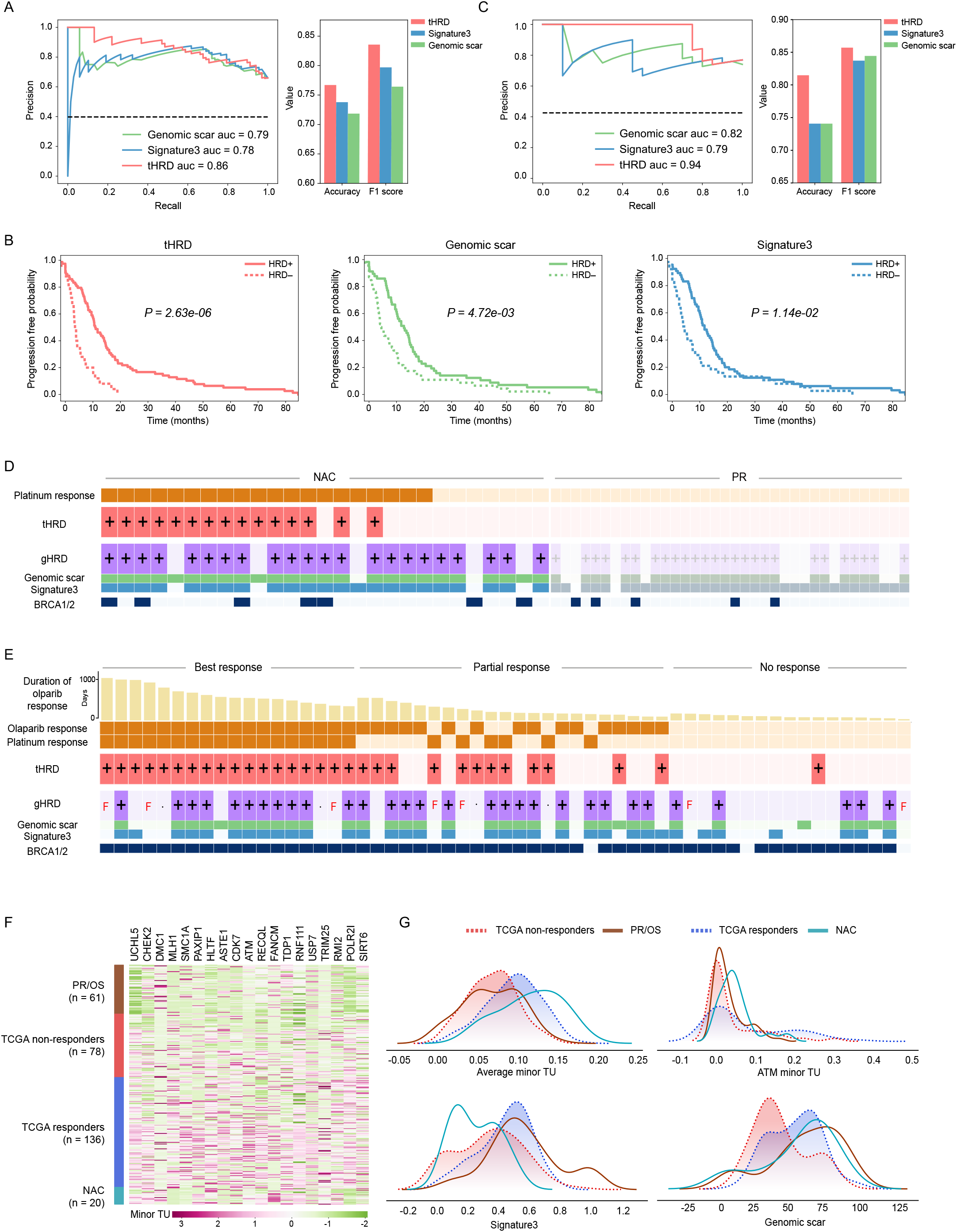
Therapeutic responses to olaparib and platinum regimens explained by tHRD. **(A-B)** HRD-based prediction of platinum sensitivity of TCGA ovarian cancer patients (n = 103). **(A)** Precision versus recall (left), and accuracy and F1 at the best accuracy (right), for tHRD, genomic scar, and signature 3. **(B)** Progression-free survival for patients partitioned by the tHRD- and gHRD-based classification. **(C)** Precision versus recall (left), and accuracy and F1 at the best accuracy (right), for tHRD, genomic scar, and signature 3 in NAC (n = 27). **(D)** Mapping of platinum responses and HRD prediction results in respective NAC (n = 27) and PR (n = 36) samples. For tHRD and signature 3, the thresholds at the best accuracy were obtained from NAC and applied to PR. For genomic scar, the previously established threshold for clinical use^58^ was employed to reduce false positives in PR. The plus signs for gHRD denote the cases in which both genomic scar and signature 3 were positive. Genetic screening results indicating the presence of germline mutations in BRCA1/2 or CHEK2 are shown at the bottom. **(E)** Mapping of olaparib/platinum responses and HRD prediction results at the best accuracy in respective OM (n = 24) and OS (n = 33) samples. Duration of olaparib responses is shown at the top. Platinum responses are marked as positive for the OM patients and negative for the OS patients. The plus signs for gHRD denote the cases in which both genomic scar and signature 3 were positive. Genome sequencing failure is depicted as ‘F’. Genetic screening results indicating the presence of germline mutations in BRCA1/2 or other genes (ATM, CHEK1, and CDK12) are shown at the bottom. **(F)** Comparison of the minor TU of selected genes (columns) across different samples (rows) from the OS/PR, TCGA, and NAC cohorts. Genes with the most significant differences between TCGA responders and non-responders were selected. **(G)** Comparison of the distribution of tHRD and gHRD metrics among the OS/PR, TCGA, and NAC groups of samples. For the average minor TU, the TU values of the minor isoform of the selected genes in (F) were averaged for each sample.

We then tested whether the tHRD model can predict responses to PARP inhibitors as well by using our data from olaparib maintenance (OM) and olaparib salvage (OS) treatment (Supplementary Table 5). The OM group (n = 24) and OS group (n = 33) data represented therapeutic responses of platinum-sensitive and platinum-resistant patients, respectively, to olaparib as second-line therapy. Whereas tHRD outperformed gHRD in predicting olaparib sensitivity overall (Fig. 6E and Supplementary Fig. 9B), we focused on the cases that were resistant to both olaparib and platinum (n = 17). Notably, both genomic scar and signature 3 were positive for 7 cases in this highly resistant group in which only one tHRD+ case was found (Fig. 6E). BRCA1/2 screening also resulted in many false positives (Fig. 6E). Demonstrating the coupling of precision to sensitivity, the gHRD methods failed to detect a few of the best responders that were sensitive to both olaparib and platinum (Fig. 6E). Among patients with partial response, sensitive to either olaparib or platinum, tHRD was positive for those with a relatively longer duration of olaparib response (Fig. 6E), thereby contributing to the clear segregation of survival curves (Supplementary Fig. 10).

To test whether the platinum resistance of the PR and OS cases can be explained by TU patterns, we partitioned TCGA samples into platinum responders and non-responders and identified key repair genes whose minor isoforms were under-expressed in the non-responders. The TU of these minor isoforms in the PR/OS samples was as low as the non-responders (Fig. 6F). The distribution of the average TU of these isoforms and that of the TU of the ATM minor isoform were similar between the PR/OS and TCGA non-responder populations (upper panel of Fig. 6G). In sharp contrast, both signature 3 and genomic scar of the PR/OS samples were distributed similarly to those of TCGA responders (lower panel of Fig. 6G).

Together with our findings about breast cancer (Supplementary Fig. 8), these findings about ovarian cancer underscore the low precision of gHRD due to confounding factors that can be either biological or technical^29^. The false positives in the PR cohort may be technical artefacts, given that most of the patients are BRCA1/2-wildtype (Fig. 6D). In contrast, our OS data highlight the influence of biological factors, considering that the olaparib users were selected based on their BRCA1/2 mutations. In this respect, the non-responders of this group are likely to involve initially imprinted genomic lesions that remain detectable after HR restoration. Given that acquired platinum resistance is common in ovarian cancer^22–25^, this raises a problem when attempting to select patient cases that will benefit from PARP inhibitors as subsequent therapy based on the gHRD signatures or BRCA1/2 mutations. In contrast, the tHRD model predicted durable responders sensitively while maintaining high precision.

The tHRD data from some of the OM and OS samples were derived from formalin-fixed paraffin-embedded (FFPE) tissues. It is known that nucleic acids are not preserved very well in FFPE tissues. Indeed, we failed DNA sequencing for some of the FFPE samples, resulting in missing data for gHRD. In contrast, our results suggest that the RNA-based tHRD assay may be applied properly to these most common sources of archived tissue material.

## CONCLUSION

PARP inhibition is a promising treatment option targeted to cancers in which DSB repair has been impaired by mechanisms such as BRCA1/2 inactivation. However, BRCA1/2 mutations comprise only a small fraction of HRD cases. Our data reinforce the notion that many HRD cases do not engage BRCA1/2 alterations. Because of the diversity of potential mechanisms, however, there has been no single unified mechanism-based assay with greater utility than the genetic screening of BRCA1/2 or HR repair gene mutations. The gHRD assays, which scan the consequences of genomic instability caused by HR defects, lack functional flexibility and suffer a high rate of false positives.

In this work, we focused on a less explored class of transcriptional aberration and found that these alterations in DNA repair-related genes prevail in breast and ovarian cancer without regard to BRCA1/2 mutations. On the basis of this widespread mechanism, we developed a novel approach that has the capacity to substitute for not only the genetic screening but also the gHRD assays. The tHRD method appears to reflect functional HR status that changes dynamically during cancer evolution, thereby minimizing biological confounding factors, in contrast to the static genome-based tests.

What remains to be tested is whether this HRD mechanism is less reversible than other mechanisms, especially under therapeutic interventions. Investigation of underlying causal factors will be helpful in this respect. Of note, these transcriptional changes involve multiple genes with sample-to-sample variations, ruling out the possibility that a single splicing factor acting in *trans* governs this somewhat complicated phenomenon; thus, genetic or epigenetic regulatory changes influencing respective genes in *cis* are likely more responsible. We employed a machine learning-based model to address this variability. In this regard, the reversion of the HRD driven in this fashion is less likely to occur than that involving single-gene coding mutations.

The utility of this method remains to be validated further beyond the multiple different contexts and settings tested in this work. Our method is expected to offer high adaptability as machine learning models can be optimized by adjusting various features and parameters. Especially, the tHRD model can be fitted directly to particular therapeutic responses instead of the universal genomic markers, as long as sufficient training data can be provided. For example, different repair genes can be used as features with variable parameters in predicting responses to different drugs in various clinical settings. In this work, we developed respective models for breast cancer and ovarian cancer. In conclusion, the tHRD-based diagnostic tests are expected to broaden the clinical utility of PARP inhibitors in various cancers.

## Data Availability

Raw sequencing data for our cohort samples have been submitted to the BioProject database under BioProject ID PRJNA700673 (reviewer access: http://www.ncbi.nlm.nih.gov/bioproject/700673).

## MATERIALS AND METHODS

### Public data collection and processing

Of 757 cases with breast adenocarcinoma from TCGA (TCGA-BRCA), we identified 644 samples with the required molecular data available for our analysis, including bam files from RNA sequencing (RNA-seq). The molecular dataset of each case included read counts normalized by counts per million (CPM), RNA-seq bam files, and somatic and germline mutation vcf files obtained from the Genomic Data Commons (GDC) portal (https://portal.gdc.cancer.gov). In addition, data regarding somatic mutation, gene expression (Illumina HiSeq pancan normalized), copy number alteration (GISTIC2.0 thresholds), methylation status (Illumina Infinium HumanMethylation450 BeadChip), genomic scar (HRD scores) and mutation signature 3 were obtained from TCGA Hub (https://tcga.xenahubs.net) and mSignatureDB (http://tardis.cgu.edu.tw/msignaturedb). Information on pathogenic or likely pathogenic germline mutations and germline mutations closely associated with two-hit events was excerpted from the supplementary data of Kuan-lin Huang et al.^1^. In addition, clinical information, including overall survival, progression-free interval, age, and tumor stage, was excerpted from the supplementary data of Liu et al.^2^.

We analyzed 324 TCGA ovarian serous cystadenocarcinoma samples (TCGA-OV) whose chemotherapy response data were available. Read counts normalized by CPM, RNA-seq bam files, and somatic and germline mutation vcf files were obtained from the GDC portal (https://portal.gdc.cancer.gov). Additional data regarding somatic mutation, gene expression (Illumina HiSeq pancan normalized), copy number alteration (GISTIC2.0 thresholds), methylation status (Illumina Infinium HumanMethylation450 BeadChip), genomic scar (HRD scores) and mutation signature 3 were obtained from TCGA Hub (https://tcga.xenahubs.net) and mSignatureDB (http://tardis.cgu.edu.tw/msignaturedb). Information on pathogenic or likely pathogenic germline mutations and germline mutations closely associated with two-hit events was excerpted from the supplementary data of Huang et al.^1^. Data for therapeutic responses to platinum chemotherapy were obtained from the supplementary data of Villalobos et al.^3^.

For the analyses of drug responses in breast cancer cell lines, RNA-seq bam flies were obtained from the GDC legacy portal (https://portal.gdc.cancer.gov/legacy-archive) while copy number alteration, methylation, and somatic mutation data were obtained from the Broad Institute Cancer Cell Line Encyclopedia (CCLE) data portal (https://portals.broadinstitute.org/ccle). Drug response data were obtained from the Release 8.2 (February 2020) of the GDSC database (https://www.cancerrxgene.org), and genetic dependency measurements from RNAi experiments were obtained from Cancer Dependency Map (DepMap) database (https://depmap.org/portal).

### Identification of BRCA1/2 mutant tumors

The presence or absence of BRCA1/2 biallelic somatic mutations and biallelic pathogenic or likely pathogenic germline mutations was examined to determine the BRCA status of TCGA-BRCA samples. We defined BRCA1/2 inactivation by biallelic somatic mutations as follows. The vcf files obtained from the GDC portal were used to identify non-silent BRCA1/2 mutation carriers that were encoded “1” by MC3 gene-level mutation annotation^4^. These cases were then investigated further to check whether their genotype was marked as an alternative homozygote (i.e., “1/1”). We also defined BRCA1/2 inactivation by biallelic germline mutations as follows. We first identified cases with pathogenic or likely pathogenic germline mutations based on the data of Huang et al.^1^. We then checked whether the pathogenic or likely pathogenic germline mutations were in alternative homozygous status (i.e., “1/1”).

We next searched for cases associated with two-hit events. First, samples with the co-occurrence of BRCA1/2 somatic or germline mutations and loss of heterozygosity (LOH) were identified. We first defined BRCA1/2 somatic inactivation coupled with LOH by identifying the carriers of MC3 gene-level non-silent somatic mutation with BRCA1/2 copy number status of “−1” (GISTIC2.0 threshold for one copy deletion). We also defined BRCA1/2 germline inactivation coupled with LOH based on pathogenic or likely pathogenic germline mutations that were reported as significantly associated with LOH (FDR < 5%)^1^ and BRCA1/2 copy number status of “-1”. Second, samples with BRCA1/2 germline with somatic mutation two-hit events were identified. Germline mutations coupled with somatic second hits were identified at the exact Poisson test P < 1×10^-5^ as previously^1^.

In addition, BRCA1/2 two copy-deleted samples were identified by looking for cases with a copy number status of “−2” (GISTIC2.0 threshold). Finally, BRCA1 epigenetic silencing was investigated based on selected probes highly associated with BRCA1 gene expression from Illumina 450K methylation data. Of all preselected probes, those localized to within 500 bp flanking the BRCA1 transcription start site were investigated. Probe cg19088651 was selected as the BRCA1 promoter methylation marker because it showed the strongest negative correlation (Pearson correlation coefficient = −0.43) with the BRCA1 gene expression level. BRCA1 promoter silencing was defined at the threshold of 0.3.

After identifying bialleic mutations, two-hit events, two-copy deletions, and epigenetic silencing, all remaining uncategorized cases were classified as BRCA1/2 active and were retained for further analysis. BRCA1/2 status of the CCLE cell lines was defined using the identical approach to that performed for TCGA-BRCA samples, based on available molecular data on copy number alteration, methylation, and somatic mutation.

### Classification of TCGA gHRD status for association with transcriptional patterns

To define differences in expression level, splicing, and transcript usage between HRD+ and HRD−, we used a stringent classification of gHRD based on using both genomic scar and signature 3. We analyzed 644 TCGA-BRCA samples. Samples were marked as gHRD+ was assigned when both genomic scar and signature 3 exceeded the median level (n = 210). Samples were marked as gHRD− when both genomic scar and signature 3 were less the median level (n = 225). These 435 samples were retained for further analysis. We analyzed 324 TCGA-OV samples in the same manner. Samples were marked as gHRD+ was assigned when both genomic scar and signature 3 exceeded the median level (n = 80). Samples were marked as gHRD− when both genomic scar and signature 3 were less the median level (n = 82).

### Identification of differentially expressed genes

Differentially expressed genes between BRCA1/2-active gHRD+ and gHRD− were identified using edgeR (v.3.28.1)^5^ with an HTseq read (v.0.6.1)^6^ count of 60,434 genes as input. Genes with less than 10 reads in less than 10 samples were filtered out. As part of the edgeR package, the quasi-likelihood F-test (glmQLFTest) was used for fitting to minimize the error rate of bulk RNA-seq methods^5^ with gHRD+/− status as a coefficient. Of our initial input of 60,434 genes, we identified 13,244 genes as differentially expressed (Benjamini-Hochberg corrected P < 0.05) with 3,672 of them repressed in the gHRD+ group. To determine whether these underexpressed genes in gHRD+ were enriched for any pathways associated with DNA repair, gene set enrichment testing using Gseapy-Enrichr (v.0.9.4)^7^ was conducted on 2,018 GO biological process terms (FDR < 5%).

### Differential splicing analysis

Differential splicing analysis was conducted on BRCA1/2-active gHRD+ (n = 169) and gHRD− (n = 219) TCGA-BRCA samples based on LeafCutter (https://davidaknowles.github.io/leafcutter/)8. Intron clustering was performed using recommended parameters, and intergroup differential splicing analysis was performed for those clusters having over 50 supporting RNA sequencing reads. Intron clusters with FDR < 10% after Benjamini-Hochberg correction were considered statistically significant and retained. Those genes with at least one intron cluster identified as significantly different were classified as differentially spliced (DS) genes. We identified 6,212 DS genes by comparing the gHRD+ and gHRD− groups and then performed functional enrichment analysis. For comparison, the DS analysis was repeated using identical parameters after randomly shuffling gHRD+ and gHRD− labels. Gene set enrichment analysis was conducted using Gseapy-Enrichr (v.0.9.4)^7^ and 2,018 biological process terms from Gene Ontology (GO) for each gene set (FDR < 5%).

### Quantification of TU and identification of aTU

Individual transcript levels were quantified by transcripts per million (TPM) through the two-pass method of StringTie (v.1.3.5)^9^ using recommended parameters. For quantification of all transcripts, including novel transcripts, we first performed the assembly step using GENCODE v29 annotation for the BRCA1/2-active TCGA-BRCA samples (n = 388) and TCGA-OV samples (n = 324), and GENCODE v19 annotation for the CCLE samples (n = 56) as reference data, to extract annotated and novel transcripts as individual gene transfer format (GTF) files for each sample. We then created a newly assembled reference by merging these GTF files of our different samples, which were used for transcript quantification. TU was calculated by first matching the gene origin of all transcripts using GffCompare (v.0.11.2)^10^ and then calculating the ratio of the respective TPM transcript to the sum of all TPM transcripts belonging to the respective matched gene.

Additional filtering was performed to identify minor transcripts with functional loss from the computed TU matrix. Major transcripts encoding proteins that are well preserved functionally and evolutionarily (defined as principal isoforms, Principals 1–5) were filtered using GENCODE v29 annotation from the APRIS (Annotating Principal Splice Isoforms) database^11^. The remaining transcripts that passed through this initial filter belonged to the Alternative 1, Alternative 2, or Not-Reported category, and were subsequently classified as minor transcripts for our use in further analysis. Next, the mean CPM of each gene was calculated to reduce noise in the minor TU matrix; the TU of genes with a mean CPM < 1 was filtered out. Additionally, standard deviation-based filtering was performed to remove TU with low intersample differences and to minimize noise in the minor TU matrix.

We used two reciprocal approaches to identifying aTU events. First, we identified minor transcripts whose TU was significantly different between gHRD+ and gHRD− samples by using the Mann-Whitney U test. The P values were adjusted based on Benjamini-Hochberg correction for TCGA-BRCA but not TCGA-OV because of small sample size. We selected the minor transcript with the highest statistical significance among multiple minor transcripts from the same gene showing significant intergroup differences (FDR < 1% for TCGA-BRCA and nominal P < 0.01 for TCGA-OV). aTU in gHRD+ was defined when the most significant minor transcript was overexpressed in gHRD+. On the other hand, aTU in gHRD− was defined when the most significant minor transcript was overexpressed in gHRD−. Second, we partitioned samples based on TU values instead of gHRD status. For each minor transcript, we selected samples whose TU was greater than the 75^th^ percentile (‘high minor TU’ group) and those whose TU was lower than the 25^th^ percentile (‘low minor TU’ group). We then compared the genomic scar scores between the two groups using the Mann-Whitney U test followed by Benjamini-Hochberg correction for TCGA-BRCA. This procedure was repeated for each minor transcript from respective genes to identify significant differences in genomic scar (FDR < 10% for TCGA-BRCA and nominal P < 0.01 for TCGA-OV). The transcript with the most significant difference in genomic scar was chosen for each gene. aTU in gHRD+ was defined when genomic scar was higher in the ‘high minor TU’ group for the most significant transcript. On the contrary, aTU in gHRD− was defined when genomic scar was higher in the ‘low minor TU’ group for the most significant transcript.

### Development of the tHRD prediction model

For the breast cancer model, we used all minor transcripts (n = 104) of DNA repair genes with aTU in gHRD+ (n = 35) as input to the random forest (RF) classifier. TCGA-BRCA samples (n = 388) were divided in a 7:3 ratio, representing the training set (n = 272) and test set (n = 116) respectively, and the TU of the 104 isoforms were used as features for prediction of gHRD status. Best parameter tuning of the RF classifier was performed using the RandomizedSearchCV module of sklearn.model. Each set of parameters went through 100 iterations, and a stratified three-fold cross-validation was performed. Mean validation accuracy was computed for each result parameter set. Parameter sets were sorted based on this mean value to identify the parameter set with the highest mean validation accuracy. The average validation accuracy for the classifier model developed with the best parameter set was 0.82. The accuracy of the model for the test set indicated 0.84. The model developed by this process was applied to the CCLE dataset to analyze the correlations of tHRD status with sensitivity to PARP inhibitors or DNA damaging drugs with the binary classification of tHRD+ or tHRD− using a threshold score of 0.5. The tHRD probability itself was used for its correlation with gene dependency (DepMap RNAi combined score).

The same procedures were repeated for the ovarian cancer model with all minor forms (n = 74) of DNA repair genes with aTU in gHRD+ genes (n = 22). The 74 TU values were used as features to develop the RF classifier model. TCGA-OV samples were divided into a training set (n = 130) and test set (n = 32). Because of its smaller sample size compared to TCGA-BRCA set, we performed a stricter best parameter tuning of the ovarian classifier to prevent overfitting. This tunning was performed by the ten-fold cross-validation option of RandomizedSearchCV. The parameter set with the highest mean validation accuracy was chosen as the best one. The accuracy of the model for the test set indicated 0.72. The model developed by this process was applied to predict platinum chemotherapy sensitivity. We employed the reannotation of the therapeutic responses of TCGA-OV samples to platinum chemotherapy^3^. Model performance was evaluated by the precision-recall curve in comparison to the gHRD metrics. The identical model was subsequently used to predict therapeutic responses to olaparib and platinum in our NAC, PR, OM, and OS cohorts.

### Clinical data collection and analysis

The NAC, PR, OM, and OS cohorts consisted of advanced-stage or recurrent ovarian cancer patients who were treated with platinum-based chemotherapy and/or olaparib from 2016 to 2021 at three hospitals (Severance Hospital, Samsung Medical Center, and Seoul National University Hospital). The NAC cohort (n = 27) included patients who were treated with neoadjuvant chemotherapy followed by interval debulking surgery. Fresh-frozen tumor samples were obtained before platinum-based chemotherapy. The PR cohort (n = 36) represented cases that showed recurrence after platinum-based chemotherapy. Fresh-frozen tumor samples were obtained after acquisition of platinum resistance (at progression). Platinum responses were regarded as sensitive with platinum-free interval (PFI) ≥ 6 months or resistant with PFI < 6 months. In the NAC cohort, 14 patients were classified as platinum-sensitive and 6 patients were platinum-resistant. All patients from the PR cohort were considered as platinum-resistant. The OM (n = 24) and OS (n = 33) cohorts, advanced-stage ovarian cancer patients treated with olaparib were enrolled for this study. The OS cohort represented patients who received olaparib as salvage therapy with platinum resistance and at least three prior lines of treatment. Most of the patients were BRCA1/2 mutants. Tumor samples, obtained before olaparib treatment, were embedded in paraffin after formalin fixation or kept fresh. The OM cohort included patients treated with olaparib for maintenance following platinum-based chemotherapy. Most of the patients were BRCA1/2 mutants. Tumor samples, obtained before the last round of platinum therapy, were embedded in paraffin after formalin fixation of kept fresh. Different criteria were applied to the OM and OS cohort when classifying the olaparib users into responders and non-responders. Complete response or partial response was considered as durable response for the OS cohort. For the OM cohort, we considered patients with progression-free survival (PFS) ≥ 12 months as responders, based on Study 19 [ClinicalTrials.gov identifier: NCT00753545] reporting the median PFS of 11.2 months for olaparib maintenance patients with BRCA1/2 mutations^12^. Clinical response was evaluated by the Response Evaluation Criteria in Solid Tumors (RECIST) 1.1. PFS was calculated from the start date of therapy to the date of progression or death. This study was approved by the institutional review board of the participating centers (Severance Hospital, Samsung Medical Center, and Seoul National University Hospital) in accordance with the Declaration of Helsinki and the International Conference of Harmonisation Good Clinical Practice guidelines (Severance Hospital, 4-2018-0342; Samsung Medical Center, 2018-10-009 and 2019-03-126; Seoul National University Hospital, 1810-035-977). Informed written consent was obtained from all patients enrolled in this study. We used a panel (CancerSCAN 3.1) to determine the presence or absence of pathogenic or likely pathogenic mutations in 15 homologous recombination repair genes (i.e., BRCA1, BRCA2, ATM, BRIP1, PALB2, RAD51C, BARD1, CDK12, CHEK1, CHEK2, FANCL, PPP2R2A, RAD51B, RAD51D, and RAD54L). Classification of variants was conducted in accordance with the principles published in the American College of Medical Genetics and Genomic standards and guidelines for interpretation of sequence variants^13^.

### Sequencing of cohort samples

Whole-genome sequencing (WGS) and whole-exome sequencing (WES) data were obtained from fresh frozen or FFPE samples. Fresh frozen tissue samples (10–30 mg) were crushed using a FastPrep-24 5G (MP Biomedicals, USA), and genomic DNA was extracted using a QIAamp DNA Mini Kit (Qiagen, Germany). For FFPE, DNA was extracted from 4 to 10 sections (4–10 μm thick) using a QIAamp DNA FFPE Tissue Kit (Qiagen). DNA concentration was measured using a Qubit dsDNA HS Assay Kit (Thermo Fisher Scientific, Waltham, MA, USA), and DNA quality was confirmed using a NanoDrop spectrophotometer (Thermo Fisher Scientific). DIN values were checked for genomic DNA integrity using Genomic DNA ScreenTape and Reagents (Agilent Technologies, Santa Clara, CA, USA). After shearing genomic DNA (500 ng) using an M220 focused-ultrasonicator (Covaris, Woburn, MA, USA), the genomic library was created using SureSelect Human All Exon V5 (Agilent Technologies, USA) or MGIEasy Universal DNA Library Prep Set (MGI Tech Co., China). Sheared DNA concentration was measured using a Qubit dsDNA HS Assay Kit (Thermo Fisher Scientific), and library fragment size was determined using a TapeStation 4200 and D1000 ScreenTape and Reagents (Agilent Technologies). DNA paired-end sequencing with 100 bp reads was performed using Illumina Hiseq2500 sequencer and Hiseq SBS kit v4 (Illumina inc., USA), or DNBSEQ-T7 sequencer and DNBSEQ-T7RS High-throughput Sequencing Kit (MGI Tech Co.).

RNA-seq data were generated from fresh frozen or FFPE samples. Fresh frozen tissue samples (10–30 mg) were crushed using a FastPrep-24 5G (MP Biomedicals), and RNA was extracted using a QIAamp DNA Mini Kit (Qiagen). For FFPE, RNA was extracted from 1 to 4 sections (4–10 μm thick) using an RNeasy FFPE mini-kit (Qiagen). RNA concentration was measured using a Qubit RNA HS Assay Kit (Thermo Fisher Scientific), and RNA quality was confirmed using a NanoDrop spectrophotometer (Thermo Fisher Scientific). RIN and DV200 values for RNA integrity were measured using a TapeStation 4200 system with RNA ScreenTape and Reagents (Agilent Technologies). For DV200 values < 40, ribosomal RNA (rRNA) was removed from RNA samples (500 ng) using an rRNA Depletion Kit (MGI Tech Co.). For DV200 values > 40, messenger RNA (mRNA) was isolated using a Dynabeads mRNA Purification Kit (Thermo Fisher Scientific). Library construction was performed using TruSeq RNA Library Prep kit v2 (Illumina inc., USA) or MGIEasy RNA Library Prep Set (MGI Tech Co.). Library concentration was measured using a Qubit dsDNA HS Assay Kit (Thermo Fisher Scientific), and library fragment size was measured using a TapeStation 4200 with D1000 ScreenTape and Reagents (Agilent Technologies). RNA paired-end sequencing with 100 bp reads was performed using Illumina Hiseq2500 sequencer and Hiseq SBS kit v4 (Illumina inc., USA), or DNBSEQ-G400 Sequencer and DNBSEQ-G400RS High-throughput Rapid Sequencing Kit (MGI Tech Co.). To align the raw reads of the PR, NAC, OS and OM cohorts, we first performed quality control using FastQC (v.0.11.9, http://www.bioinformatics.babraham.ac.uk/projects/fastqc), followed by trimming by Trim Galore (v.1.18, https://www.bioinformatics.babraham.ac.uk/projects/trim_galore). Alignments were performed using a two-pass strategy with STAR (v.2.7.1a)^14^ with recommended options.

### Estimation of signature 3 and genomic scar of cohort samples

To estimate the signature 3 contribution of the PR, NAC, OS, and OM samples, single nucleotide variant (SNV) calling was performed using Mutect2 (v.2.2)^15^ with recommended filtering steps and default parameters. For the PR and OS cohort, Mutect2 matched-normal mode with default parameters was used with WES bam files as input for SNV calling, whereas Mutect2 tumor-only mode with default parameters was used for SNV calling from WGS bam files as input for the NAC and OM cohort. For the CCLE samples, we obtained the calling data for somatic mutations from the Broad Institute CCLE portal (https://portals.broadinstitute.org/ccle). Next, we determined the mutation signature of each SNV using the R package deconstructSigs (v.1.8.0)^16^. Variant information in the vcf files was transformed into the recommended input file format by using the vcf.to.sigs.input method provided by the software package. To assess signature contributions, fitting was performed for the previously reported set of mutational signatures for each cancer type (i.e., signatures 1, 3, and 5 in ovarian cancer and signatures 1, 2, 3, 5, 6, 8, 10, 13, 17, 18, 20, 26, and 30 in breast cancer) using the Catalogue of Somatic Mutations in Cancer (COSMIC) database (https://cancer.sanger.ac.uk/cosmic/signatures_v2).

Genomic scar was estimated by determining copy number alterations in the WES or WGS data using sequenza-utils (v.3.0.0)^17^, based on which LOH, large scale transitions, and the number of telomeric allelic imbalances were estimated using the scarHRD R package (Rpackage v.0.1.1)^18^. The sum of these values served as the genomic scar score^19–21^. We calculated genome-wide GC content values by using the gc_wiggle program with recommended parameters. We then processed the genomic bam files with the GC content values by using the bam2seqz program with recommended parameters to convert to a seqz file that contains information on copy number alterations. The process to determine copy number alterations were identical to SNV calling with Mutect2. The matched-normal mode was used for the PR and OS WES bam files. The tumor-only mode was used for the NAC WGS bam files with the Pan-Cancer Analysis of Whole Genomes (PCAWG) normal WGS bam file as non-matching normal samples. Because the seqz file of each cohort obtained through sequenza-utils was split for each chromosome, a single seqz file was merged per sample. Genomic scar was calculated using the scarHRD R package (Rpackage v0.1.1)^18^ and used as the input seqz file. gHRD status was determined by using the median of signature 3 and genomic scar scores in respective cohorts.

### Functional assays

Primary breast cancer (PD:tHRD+ and PD:tHRD−) cells were isolated from tumor tissues of a patient-derived xenograft (PDX) model. Fresh tumor tissues were minced into 1–2 mm pieces using sterile scissors, scalpel, and forceps. For tissue digestion, the tissue pieces were incubated in RPMI 1640 medium (Hyclone, USA) containing 5% FBS (Hyclone), 1% penicillin/streptomycin (Hyclone), 20 µg/ml of collagenase Type Ⅲ (Sigma Aldrich, USA), and 840 ng/ml Amphotericin B (Gibco, USA) for 6 hrs at 37°C. The digested tissue pieces were washed with media three times. The primary cells were cultured in RPMI 1640 medium containing 5% FBS, 1% penicillin/streptomycin, hEGF (20 ng/ml, Gibco), hydrocortisone (4 μg/ml, Sigma Aldrich, and transferrin (4 μg/ml, Sigma Aldrich). Depletion of mouse stromal cells was performed using Mouse Depletion Kit (Miltenybiotec, Germany) according to the manufacturer’s instructions.

For immunocytochemistry, the PD:tHRD+ and PD:tHRD− cells were cultured in a slide-covered dish and treated with Olaparib (2 μM, Sigma) for 48 hrs. The cells were fixed with 4% formaldehyde and permeabilized with 0.1% Triton X-100. Sample slides were labeled with primary antibodies for Rad51 (1:500; Abcam, USA) or γ-H2AX (1:1000, Abcam) and then stained with the corresponding Alexa Fluor 488-conjugated secondary antibody (Invitrogen, Carlsbad, CA, USA) for 1 h at room temperature. DAPI was used for nuclear staining. The cells were then mounted with Aqua-Poly-Mount mounting medium and imaged using an LSM710 confocal microscope (Carl Zeiss AG, Oberkochen, Germany) under 400X magnification. 20 cells from each sample were counted to calculate the nuclear Rad51 and γ-H2AX foci.

For HR activity assays, the PD:tHRD+ and PD:tHRD− cells were pretreated with control media or with Olaparib (2 μM). After 12–16 hrs, the cells were transfected with circular pDR-GFP (Addgene 26475) using Lipofectamine 2000 as recommended by the manufacturer (Invitrogen). After 24 hrs, the cells were additionally transfected with the I-SceI-producing plasmid pCBASceI (Addgene 26477). Flow cytometry analysis was performed 24 hrs later with AccuriFlow Cytometry (BD Biosciences, USA). The GFP-positive cells were quantified by the FlowJo program, and the average HR frequency was measured from three independent samples.

We performed Western blotting as follows. After Olaparib treatment for 48 hrs, protein samples were prepared and loaded at 10–50 µg of total protein per lane. The blot was probed with anti-BCL2 (1:1000, Cell Signaling Technology, USA), anti-BAX (1:1000, Cell Signaling), anti-phospho-p53 (Ser-15 or −392) (1:1000, Cell Signaling), and anti-β-actin (1:1000, Santa Cruz Biotechnology, USA) antibodies. The relative densities of bands were analyzed with the NIIH ImageJ 1.47v software. For the alamarBlue® cell viability assay, cells were cultured in a 96-well plate and treated with 2 mM Olaparib. At 24, 48, and 72 hrs post-treatment, 1/10th volume of alamarBlue® reagent (Invitrogen) was added directly to the culture media. The alamarBlue® assay was performed according to the manufacturer’s instructions. Viable cells were detected by fluorescence measurements using a microplate fluorescence spectrophotometer (GenTeks Biosciences, Inc., San-Chong, Taipei).

## SUPPLEMENTARY FIGURE LEGENDS

**Supplementary Figure 1.**
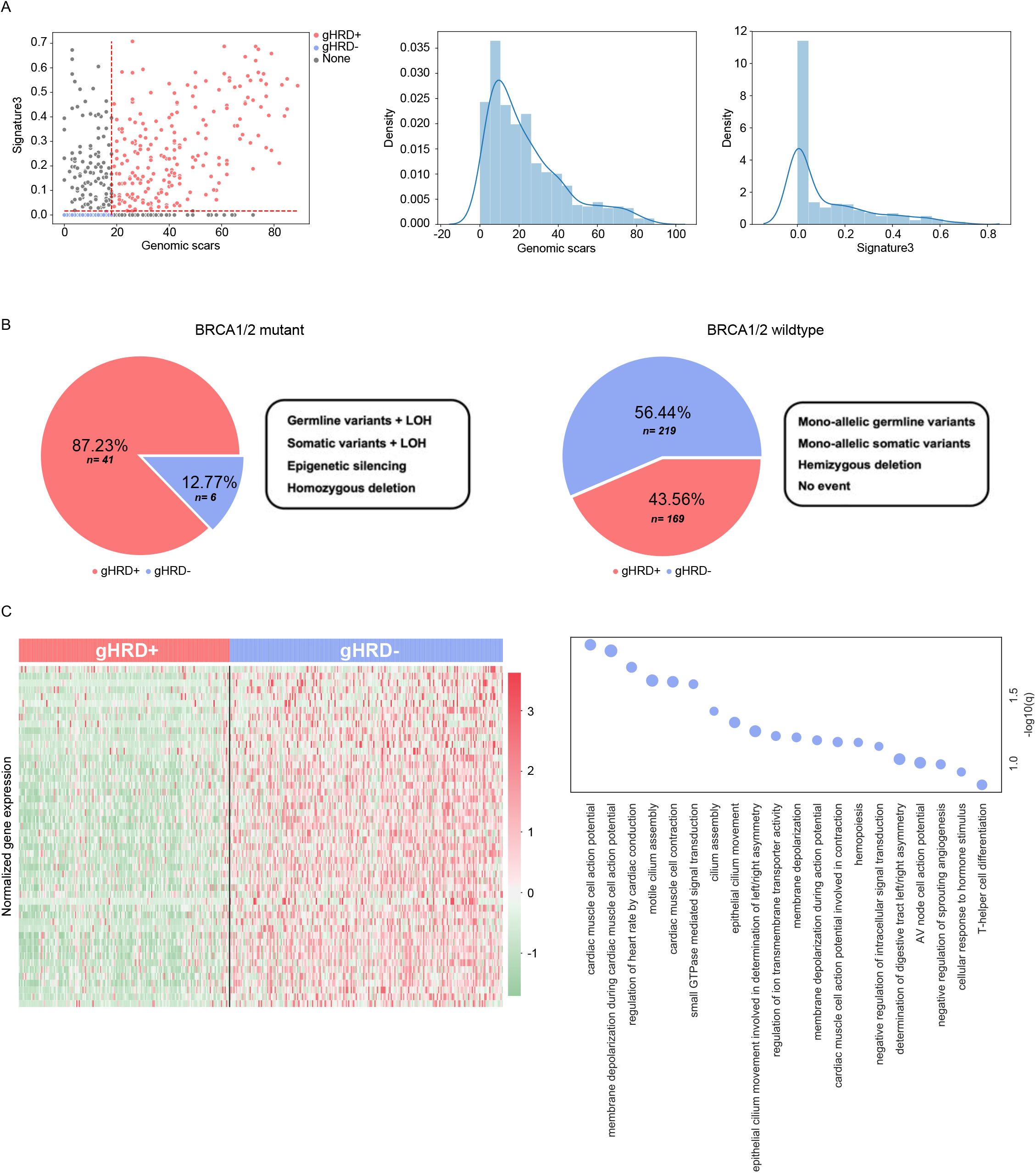
gHRD distribution and corresponding gene expression in breast cancer. **(A)** Distribution of signature 3 and genomic scar in TCGA samples (n = 644). In this analysis, gHRD+ and gHRD− were defined when both signature 3 and genomic scar were above or below the median denoted by the dotted lines of the leftmost scatter plot. **(B)** Proportion of the gHRD+ samples in BRCA1/2 mutation and wildtype groups. To define BRCA1/2 mutants, we included germline/somatic variation coupled with loss of heterozygosity (LOH), epigenetic silencing by promoter methylation, and homozygous deletion (left pie chart). Mono-allelic germline or somatic variants and hemizygous deletions were regarded as BRCA1/2 wildtype (right pie chart). **(C)** Expression level heatmap (left) and functional enrichment plot (right) for the genes down-regulated in the gHRD+ tumors.

**Supplementary Figure 2.**
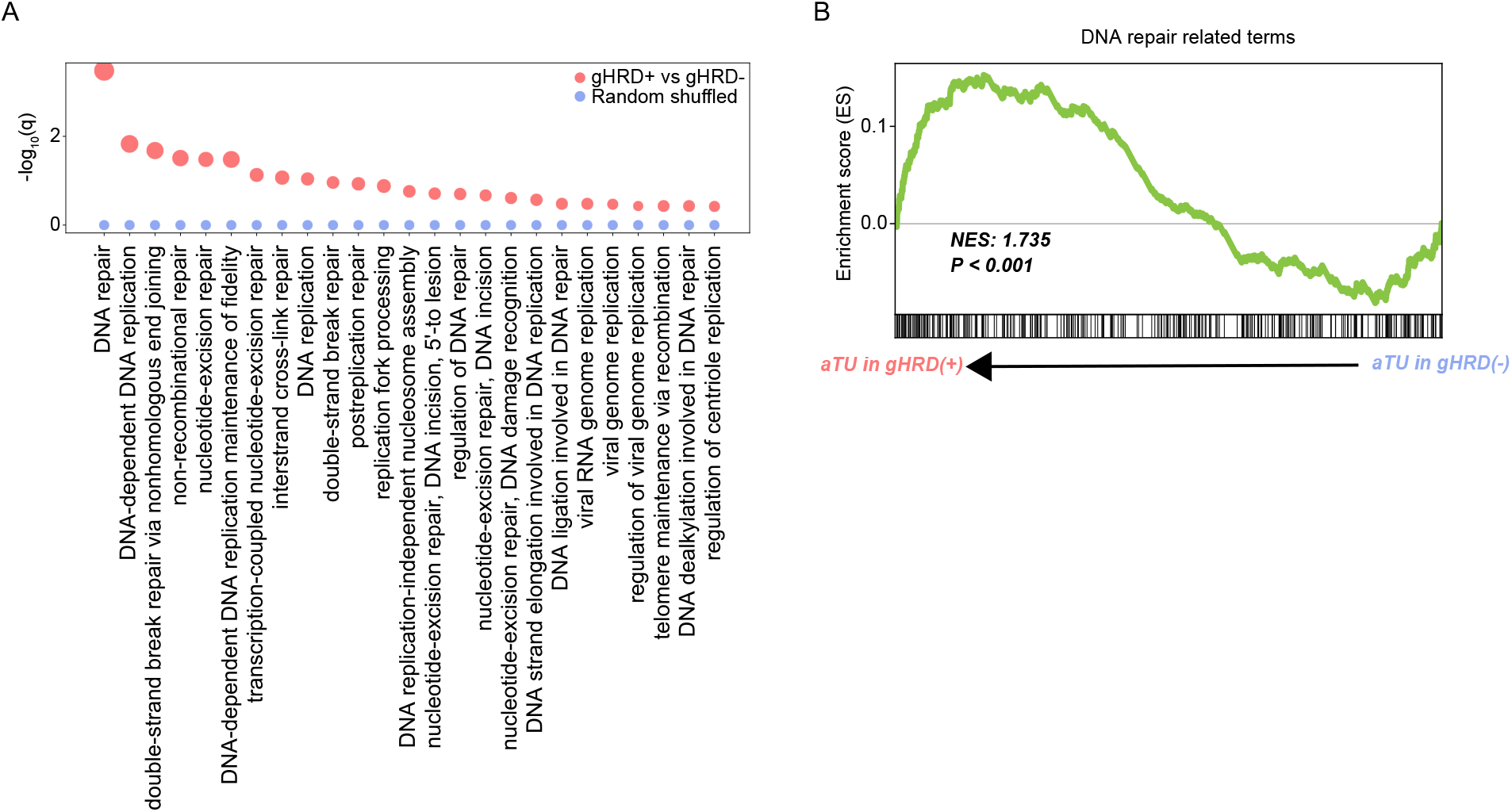
HRD-associated alterations in splicing of DNA repair genes. Genes enriched for differential RNA splicing between the gHRD+ and gHRD− tumors. The statistical significance of overrepresentation of the genes related to DNA repair, recombination, and replication is compared between sample groupings by gHRD status versus by shuffled labels. **(B)** Gene Set Enrichment Analysis (http://www.gsea-msigdb.org) plot for DNA repair-related terms in association with aTU. Genes are sorted according to differential minor TU between gHRD+ and gHRD− such that genes with higher minor TU in gHRD+ are found on the left and those with higher minor TU in gHRD− are found on the right. For each gene, the most differential isoform in either direction has been selected. Black bars depict the position of genes belonging to DNA repair-related terms. Normalized enrichment score (NES) and its P value for these terms are shown.

**Supplementary Figure 3.**
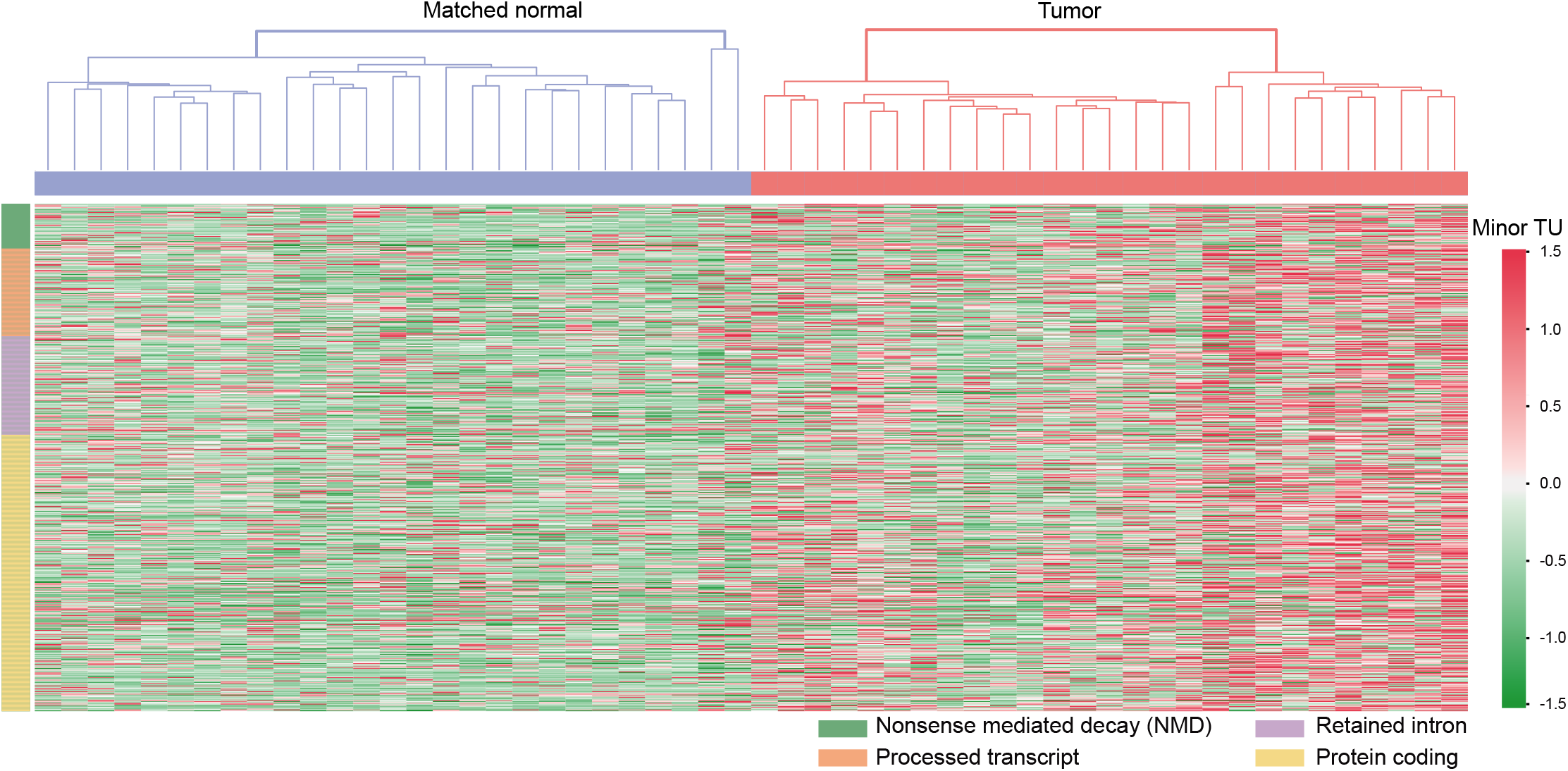
Differential minor TU between tumor and matched normal tissues. Heatmap comparing minor TU levels of the aTU genes (rows) between the gHRD+ tumor and matched normal samples (columns). Shown are 49 samples with matched normal transcriptome data available. Annotation categories that represent the functional consequences or alternative events by minor isoform transcription are provided at the bottom.

**Supplementary Figure 4.**
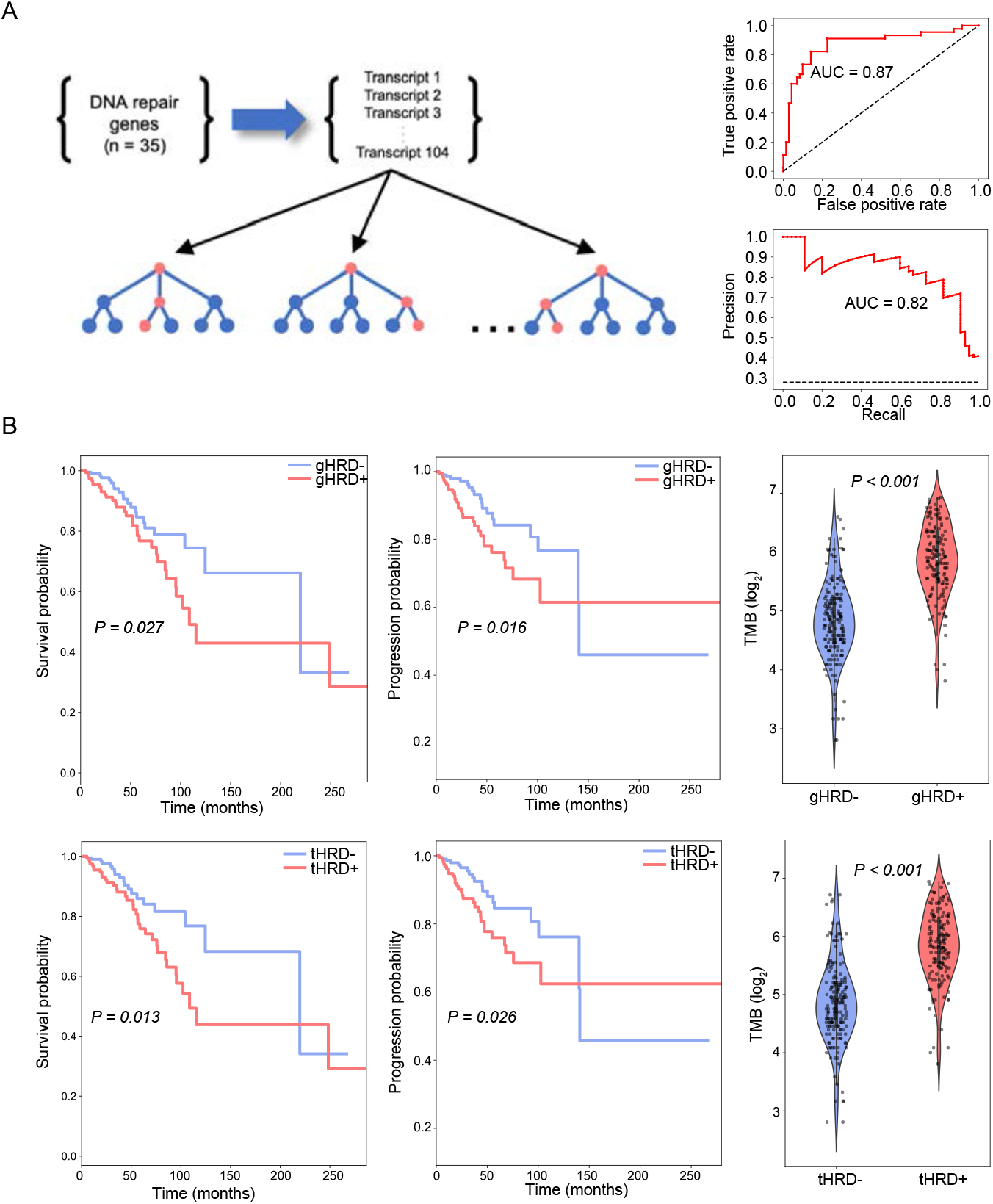
tHRD model in breast cancer. **(A)** For the random forest model, we selected 35 DNA repair genes that showed aTU in breast cancer and used the TU level of their 104 minor transcripts as input to the prediction model. Performance of the aTU-based classifier in predicting gHRD was measured. **(B)** Comparison of tHRD and gHRD in terms of patient prognosis and tumor mutation burden (TMB). Overall survival (left), progression-free survival (middle), and TMB (right) are compared between the gHRD classification (upper) and tHRD prediction (lower).

**Supplementary Figure 5.**
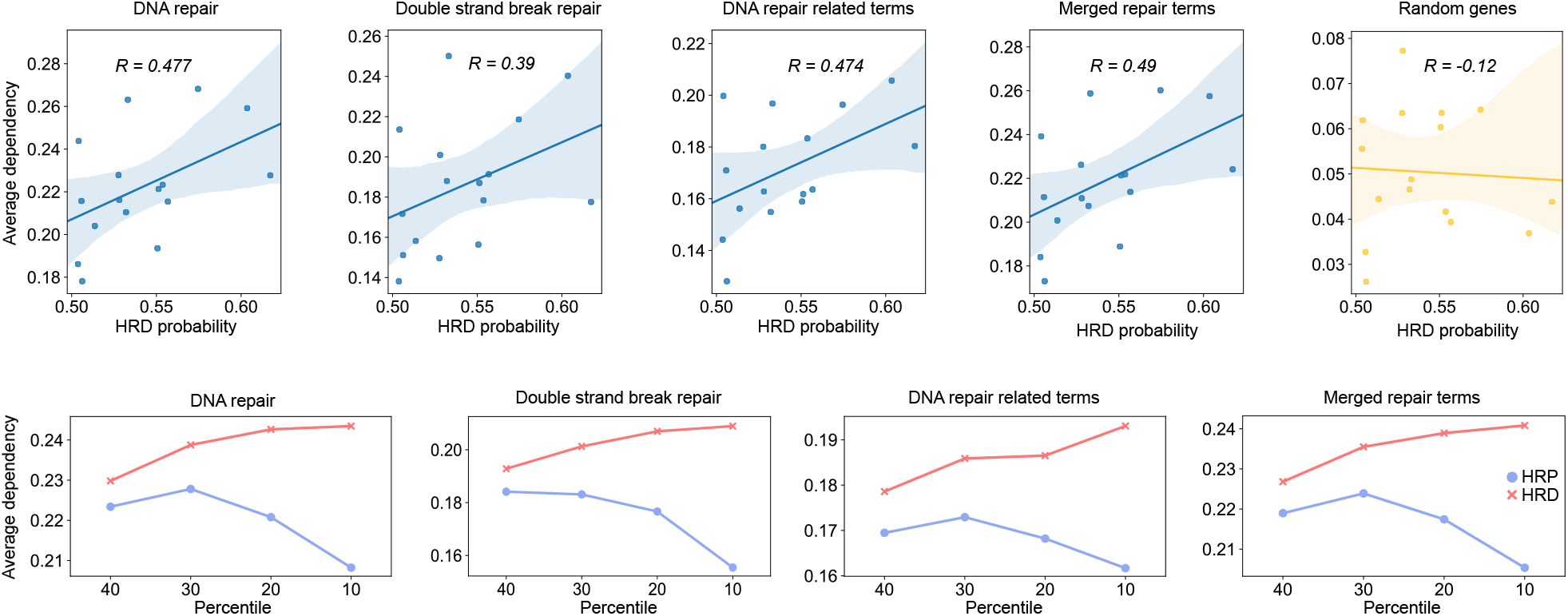
Functional dependency of tHRD+ cell lines on DNA repair-related genes. Scatter plots of the tHRD score of tHRD+ cell lines (x-axis) versus the average dependency on relevant genes (y-axis) (upper), and graphs comparing the average dependency of the tHRD+ versus tHRD− cell lines on relevant genes according to the percentile of the tHRD score (lower). Dependency data were obtained from the Cancer Dependency Map database (https://depmap.org/).

**Supplementary Figure 6.**
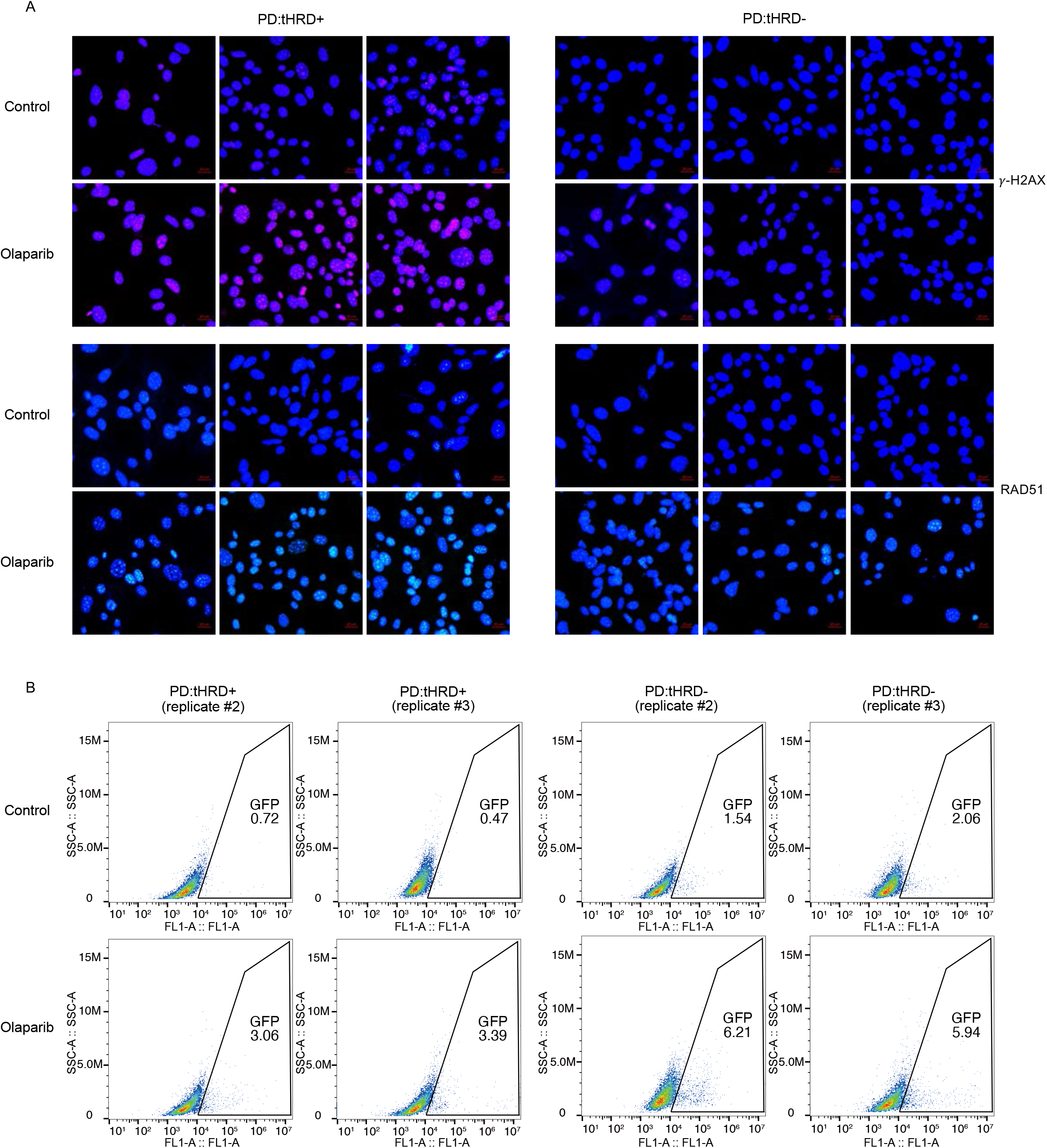
Functional HR assay of tHRD+ and tHRD− PD cells. **(A)** Additional immunofluorescence staining images for γ-H2AX and RAD51 in PD:tHRD+ and PD:tHRD− before and after olaparib treatment. Representative images are provided in Fig. 4B. **(B)** Scatter plots from replicate experiments to measure the rate of HR repair based on the DR-GFP/I-*Sce*I assay in PD:tHRD+ and PD:tHRD− before and after olaparib treatment. GFP-positive cells in the marked zones indicate the proportion of cells that underwent HR-mediated DSB repair of GFP reporter plasmids. Representative plots are provided in Fig. 4C.

**Supplementary Figure 7.**
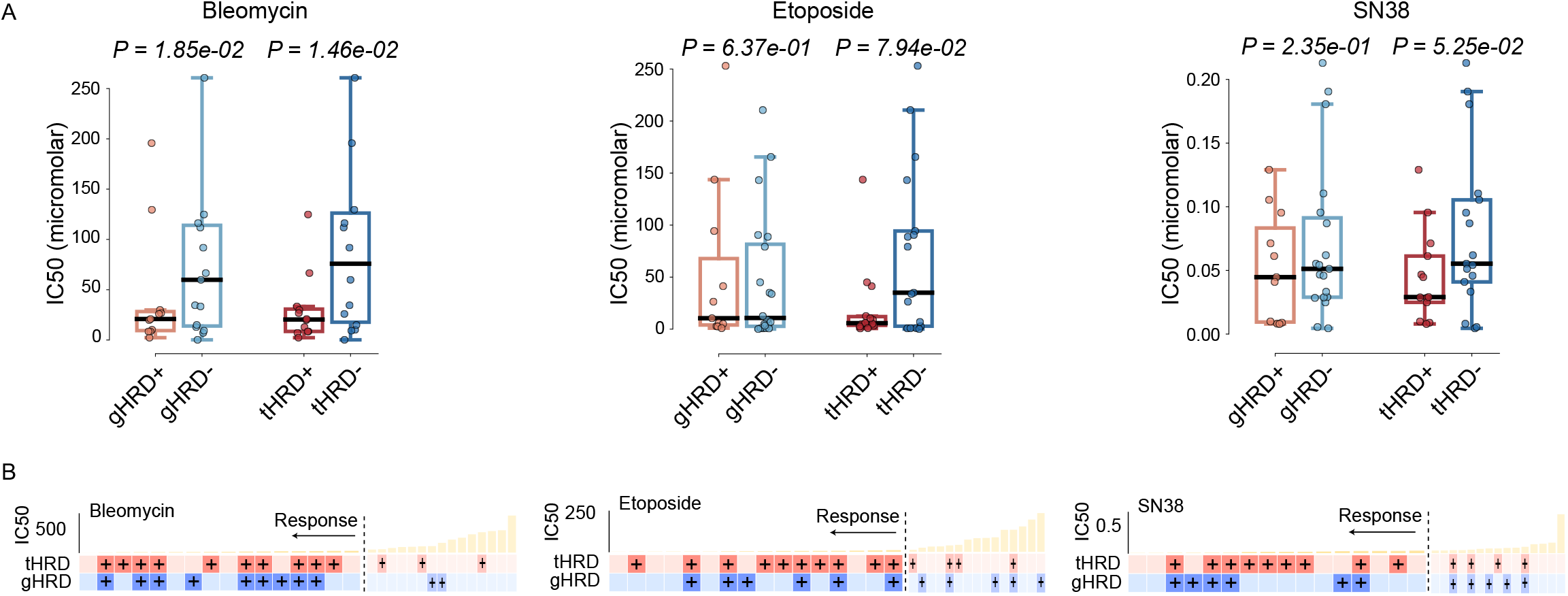
Responses to DNA damaging agents explained by tHRD. **(A)** Comparison of IC_50_ between tHRD+/tHRD− versus gHRD+/gHRD− in breast cancer cell lines. Response data were obtained from the GDSC database. Our tHRD detection model was applied to the RNA sequencing data of the cell lines to classify them as tHRD+ and tHRD−. **(B)** Mapping of drug sensitivity and HRD prediction results in respective cell lines. Below the plot of IC_50_ values are the results of our tHRD model and signature 3-based gHRD assay for the corresponding cell lines. The plus signs depict HRD+ predictions. gHRD was considered positive above the 75^th^ percentile of signature 3. IC_50_ values lower than the median were regarded as positive responses to the given drug.

**Supplementary Figure 8.**
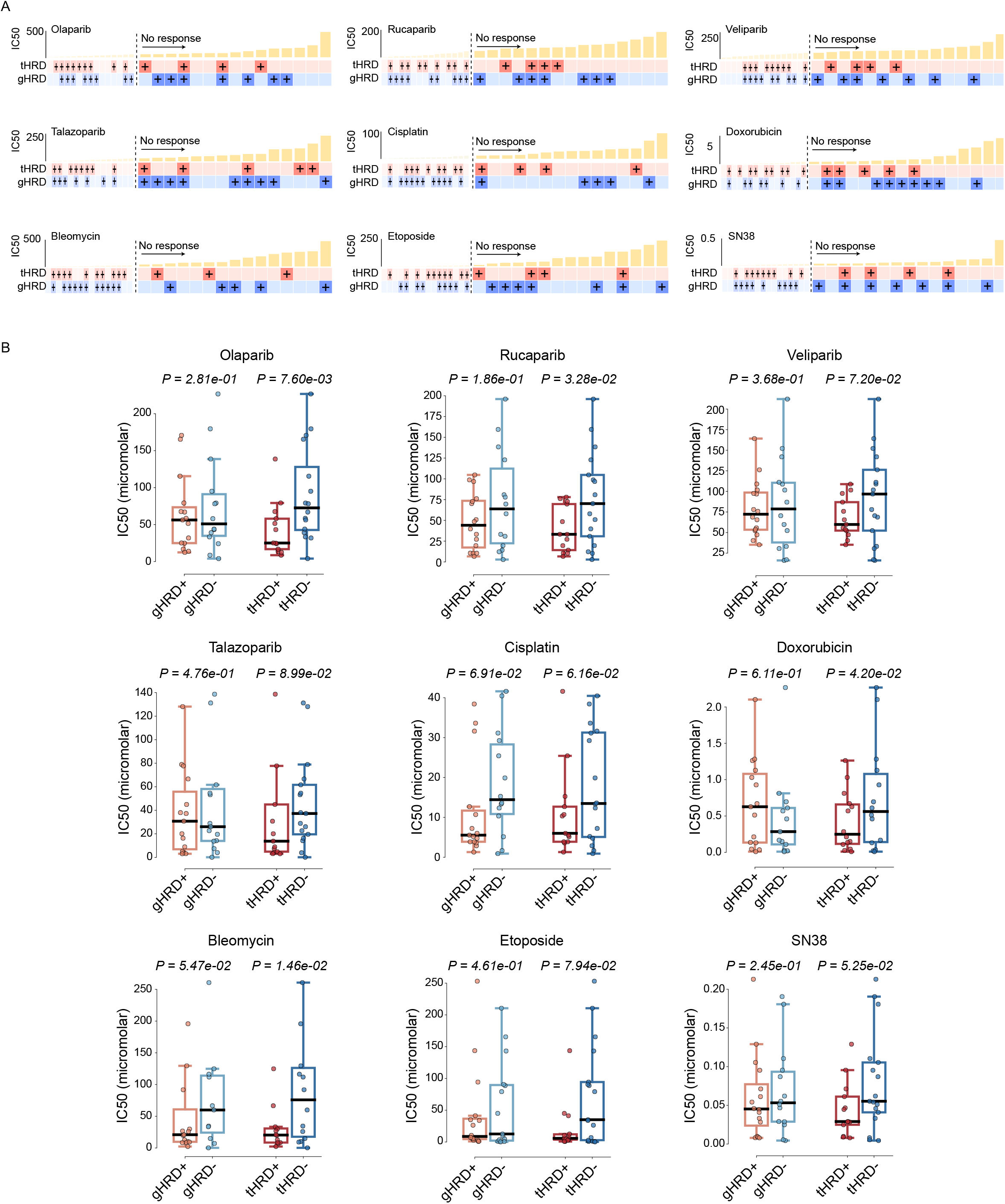
Drug responses at a lower gHRD threshold. **(A)** Same plots as Fig. 5B and Supplementary Fig. 7B after lowering the threshold of gHRD (from the 75^th^ to 50^th^ percentile) **(B)** Same plots as Fig. 5A and Supplementary Fig. 7A after lowering the threshold of gHRD (from the 75^th^ to 50^th^ percentile).

**Supplementary Figure 9.**
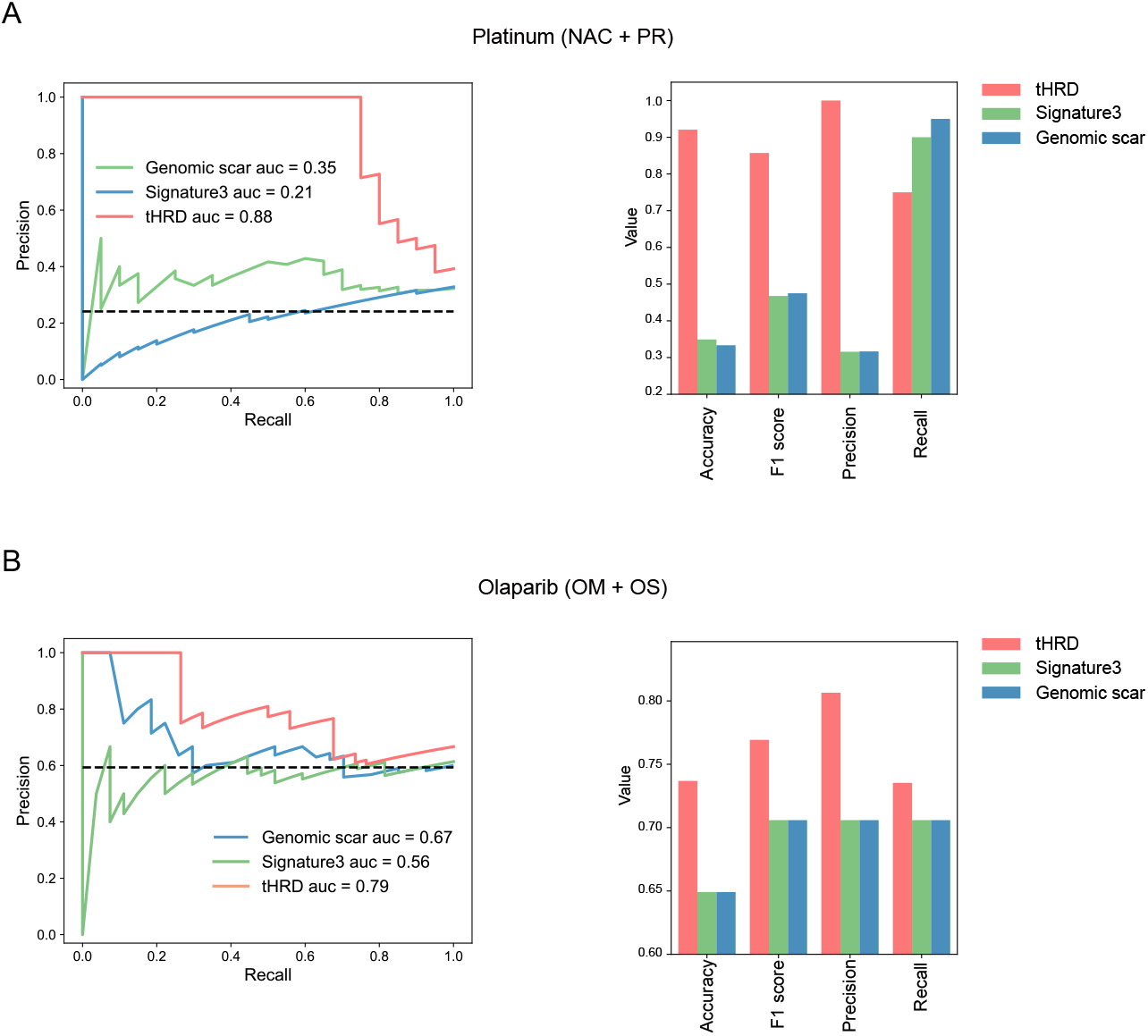
Performance of tHRD and gHRD in predicting sensitivity to platinum or olaparib. Precision versus recall (left), and accuracy, F1, precision, and recall at the best accuracy (right), for tHRD, genomic scar, and signature 3 in **(A)** NAC and PR combined (n = 63) and **(B)** OM and OS combined (n = 57).

**Supplementary Figure 10.**
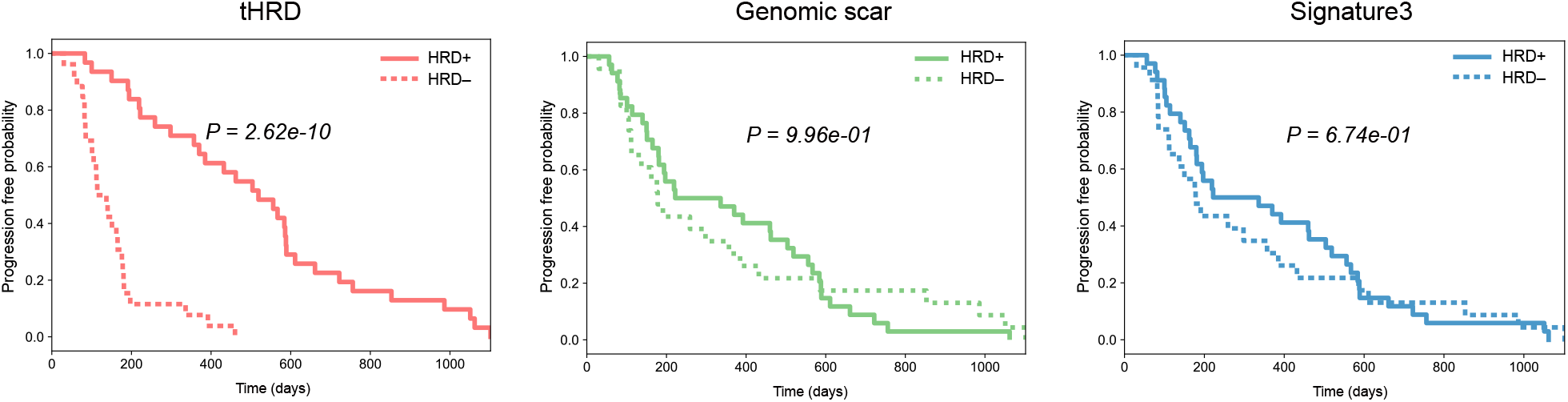
Patient survival explained by tHRD and gHRD in response to olaparib. Progression-free survival of the OM and OS patients (n = 57) segregated by the tHRD- and gHRD-based classification. Duration of olaparib responses plotted in Fig. 6D was used.

